# Estimation of Life’s Essential 8 Score with Incomplete Data of Individual Metrics

**DOI:** 10.1101/2023.03.03.23286786

**Authors:** Yi Zheng, Tianyi Huang, Marta Guasch-Ferre, Jaime Hart, Francine Laden, Jorge Chavarro, Eric Rimm, Brent Coull, Hui Hu

## Abstract

**Background:** The American Heart Association’s Life’s Essential 8 (LE8) is an updated construct of cardiovascular health (CVH), including blood pressure, lipids, glucose, body mass index, nicotine exposure, diet, physical activity, and sleep health. It is challenging to simultaneously measure all eight metrics at multiple time points in most research and clinical settings, hindering the use of LE8 to assess individuals’ overall CVH trajectories over time.

**Methods and Results:** We obtained data from 5,588 participants in the Nurses’ Health Studies (NHS, NHSII) and Health Professional’s Follow-up Study (HPFS), and 27,194 participants in the 2005-2016 National Health and Nutrition Examination Survey (NHANES) with all eight metrics available. Individuals’ overall cardiovascular health (CVH) was determined by LE8 score (0-100). CVH-related factors that are routinely collected in many settings (i.e., demographics, BMI, smoking, hypertension, hypercholesterolemia, and diabetes) were included as predictors in the base models of LE8 score, and subsequent models further included less frequently measured factors (i.e., physical activity, diet, blood pressure, and sleep health). Gradient boosting decision trees were trained with hyper-parameters tuned by cross-validations. The base models trained using NHS, NHSII, and HPFS had validated root mean squared errors (RMSEs) of 8.06 (internal) and 16.72 (external). Models with additional predictors further improved performance. Consistent results were observed in models trained using NHANES. The predicted CVH scores can generate consistent effect estimates in associational studies as the observed CVH scores.

**Conclusions:** CVH-related factors routinely measured in many settings can be used to accurately estimate individuals’ overall CVH when LE8 metrics are incomplete.

**Clinical Perspective:** *What Is New?:* Life’s Essential 8 (LE8) has great potential to assess and promote cardiovascular health (CVH) across life course, however, it is challenging to simultaneously collect all eight metrics at multiple time points in most research and clinical settings. We demonstrated that CVH-related factors routinely collected in many research and clinical settings can be used to accurately estimate individuals’ overall CVH across time even when LE8 metrics are incomplete.

*What Are the Clinical Implications?:* The approach introduced in this study provides a cost-effective and feasible way to estimate individuals’ overall CVH. It can be used to track individuals’ CVH trajectories in clinical settings.

## Introduction

Cardiovascular disease (CVD) is the top cause of death both in the United States (US) and globally.^1^ It is estimated that 80% of CVD is preventable.^2^ Conventional CVD prevention strategies emphasize the optimizations of classical risk factors such as blood pressure and lipids. However, it is challenging to communicate CVD risk to young individuals with a low absolute 10-year CVD risk. To address this, the American Heart Association (AHA) introduced the Life’s Simple 7 (LS7) in 2010, to assess and promote cardiovascular health (CVH),^3^ which anchors CVD prevention in health rather than disease to prompt attention to primordial prevention across life course.^4^ The AHA defined ideal CVH based on seven metrics (LS7), including blood pressure, total cholesterol, glucose, body mass index (BMI), cigarette smoking, diet, and physical activity.^3^ To better account for factors predictive of CVH, the AHA recently introduced Life’s Essential 8 (LE8), an updated construct of CVH with revised quantitative assessment of the 7 existing metrics as well as one new metric focusing on sleep health.^5^ Previous studies have shown that CVH is not only associated with CVD,^6,7^ but also non-CVD outcomes such as cancer,^8^ cognitive impairment,^9^ depression,^10^ and all-cause mortality.^11^

In 2016, the AHA announced an ambitious initiative, One Brave Idea,^12^ with the goal to end coronary heart disease and its consequences. An interim target called “50×50×50” was proposed in 2018, with the goal of achieving ideal CVH among “≥50% segments of the population ≤50 years old by 2050 or sooner”.^13^ Previous estimates based on LS7 showed that the prevalence of ideal CVH in the US population is around 50% at 10 years of age and declines to less than 10% by 50 years of age.^14,15^ Similarly, recent estimates based on LE8 showed that compared with individuals aged 12-19 years, the mean CVH score is 13.9% lower among those aged 40-64 years.^16^ Therefore, it is important to understand population-level CVH trajectories and identify factors contributing to different CVH trajectories to promote and preserve CVH. However, to date, population-level CVH estimates are mainly cross-sectional.^14,17–22^ Very few studies have examined individuals’ CVH trajectories over time.^23–28^ Among these existing studies, CVH trajectories were determined based on either CVH status sparsely measured over time (e.g., 3 time points in ≥10 years),^23–25^ or modified versions of LS7 where not all CVH metrics were considered.^25–28^ This is mainly due to the challenges of having all CVH metrics simultaneously measured at multiple time points, which substantially hindered the adoption of LE8 to promote and preserve CVH across life course. It remains unclear regarding the performance of a subset of LE8 metrics in estimating overall CVH defined by the full LE8 metrics.

To address this limitation, leveraging data from the Nurses’ Health Study (NHS), the Nurses’ Health Study II (NHSII), the Health Professional’s Follow-up Study (HPFS), and the 2005-2016 National Health and Nutrition Examination Survey (NHANES), we developed and validated models to estimate individuals’ overall CVH using CVH-related factors that are routinely collected in many research and clinical settings to enable longitudinal assessment of CVH trajectories even when not all eight CVH metrics are available simultaneously.

## Methods

### Study Population

We obtained data from three large nationwide prospective cohorts in the U.S., including NHS and NHSII, with 121,700 and 116,429 female registered nurses recruited in 1976 and 1989, respectively, as well as HPFS, with 51,529 male health professionals recruited in 1986. We also obtained data from the 2005-2016 NHANES, a complex survey with nationally representative samples of noninstitutionalized U.S. adults. A total of 5,588 participants from the cohorts (i.e., 4,114 from NHS, 676 from NHSII, and 798 from HPFS) and 27,194 participants aged 18 and older from the 2005-2016 NHANES with all eight CVH metrics measured.

### Assessment of Individual CVH Metrics

Blood samples were collected in NHS in 1989-1990 (n=32,826), NHSII in 1996-1999 (n=29,611), and HPFS in 1993-1995 (n=18,159). Among them, a total of 5,030, 785, and 1,388 participants in NHS, NHSII, and HPFS, respectively, had both hemoglobin A1c (HbA1c) and blood lipids measured in the same blood sample. In the 2005-2016 NHANES, HbA1c was measured in whole blood biospecimen using chromatogram, and blood lipids was measured in serum sample using an enzymatic assay.^29^ Measures of the other six metrics (i.e., BMI, nicotine exposure, blood pressure, diet, physical activity, and sleep health) were obtained in NHS, NHSII, and HPFS based on self-reports from questionnaires closest to blood sample collections (Table S1). Previous validation studies showed that these self-reported measures are highly accurate.^30–44^ Participants in NHS, NHSII, HPFS cohorts were asked about their typical systolic and diastolic blood pressure (i.e., systolic pressure: <105, 105-114, 115-124, 125-134, 135-144, 145-154, 155-164, 165-174, and ≥175 mmHg; diastolic pressure: <65, 65-74, 75-84, 85-89, 90-94, 95-104, and ≥105 mmHg). In NHANES, participants’ blood pressures were consecutively measured multiple times with at least 5 minutes of break between measurements, and the average blood pressure was used. Self-reported history of medications on hypertension (i.e., thiazide diuretics, alpha blockers, beta blockers, calcium channel blockers, angiotensin-converting enzyme inhibitors, Lasix, and other anti-hypertensive medications), diabetes (i.e., insulin, and oral hypoglycemic medications), and hypercholesterolemia (i.e., statin and other cholesterol-lowering medications) was used to determine controlled treatments in both the cohorts and NHANES. BMI was calculated based on self-reported weight and height in the cohorts, while in NHANES, weight and height were measured by physical examinations. Nicotine exposure was assessed by self-reports in both the cohorts and NHANES. In the cohorts, physical activity was computed by summing up the metabolic equivalent of task (MET) hours of each individual activity per week according to corresponding MET score and self-reported hours of the activity.^43,45^ In NHANES, physical activity was determined based on self-reported frequency and duration of moderate- and vigorous-intensity leisure time activities, with 4 MET scores assigned to each minute of moderate activities and 8 MET scores assigned to each minute of vigorous activities. Diet was assessed by a >130-item validated food frequency questionnaire in the cohorts,^36,38–42^ and by 24-hour dietary recall in NHANES. Sleep health was assessed by self-reported average sleep hours during a 24-hours period in both NHANES and the cohorts. The eight individual CVH metrics (i.e., blood pressure, lipids, glucose, BMI, nicotine exposure, diet, physical activity, and sleep health) were scored with a range from 0 to 100. Table 1 shows the detailed scoring criteria for each metric. Specifically, we used the same criteria recommended by the AHA to assess blood lipids, nicotine exposure, and physical activity.^3,5^ For blood pressure, we used a slightly different sets of cut points because (1) these were the cut-points used in the questionnaires for NHS, NHSII, and HPFS, (2) although the American College of Cardiology (ACC)/AHA hypertension clinical practice guideline set 130/80 mmHg as the cut point for hypertension diagnosis,^46^ the International Society of Hypertension Global Hypertension Practice Guidelines set average day time ambulatory blood pressures or home blood pressure >135/85 mmHg as the criteria for hypertension diagnosis,^47^ and (3) it has been shown that any blood pressure over 115/75 increases the risk of CVD.^48–50^ HbA1c was used to assess the glucose metric since fasting blood glucose was not collected in NHS, NHSII, and HPFS. Moreover, HbA1c test is recommended and clinically used to detect diabetes with high validity and cost-effectiveness,^51,52^ and widely used in other studies to assess CVH.^53–56^ In addition, alternative healthy eating index 2010 (AHEI-2010) was used to measure adherence to a healthy diet pattern based on foods and nutrients that are predictive of chronic disease risk and has been used to assess diet-disease associations in many published studies.^57–59^ Percentiles of AHEI-2010 scores were used to assess status of diet.

**Table 1.** Scoring criteria of CVH metrics based on Life’s Essential 8.

| **Metric** | **Points and Criteria** |
| --- | --- |
| **Blood pressure** | 100: SBP<115mmHg and DBP<75mmHg<br>75: SBP 115-124mmHg and DBP<75mmHg<br>50: SBP 125-134mmHg or DBP: 75-84mmHg<br>25: SBP 135-154mmHg or DBP 85-94mmHg<br>0: SBP ≥155mmHg or DBP≥95mmHg<br>(Subtract 20 points if treated level) |
| **HbA1c** | 100: No history of diabetes and HbA1c<5.7%<br>60: No diabetes and HbA1c 5.7-6.4%<br>40: Diabetes with HbA1c<7.0%<br>30: Diabetes with HbA1c 7.0-7.9%<br>20: Diabetes with HbA1c 8.0-8.9%<br>10: Diabetes with HbA1c 9.0-9.9%<br>0: Diabetes with HbA1c≥10.0% |
| **Blood lipids** | 100: Non-HDL cholesterol<130 mg/dL<br>60: Non-HDL cholesterol 130-159 mg/dL<br>40: Non-HDL cholesterol 160-189 mg/dL<br>20: Non-HDL cholesterol 190-219 mg/dL<br>0: Non-HDL cholesterol ≥220 mg/dL<br>(Subtract 20 points if treated level) |
| **Nicotine exposure** | 100: Never smoker<br>75: Former smoker, quit ≥5 year<br>50: Former smoker, quit 1-5 year<br>25: Former smoker, quit<1 year<br>0: Current smoker<br>(Subtract 20 points if living with active indoor smoker in home) |
| **BMI** | 100: <25kg/m<sup>2</sup><br>70: 25.0-29.9kg/m<sup>2</sup><br>30: 30.0-34.9kg/m<sup>2</sup><br>15: 35.0-39.9kg/m<sup>2</sup><br>0: ≥40.0kg/m<sup>2</sup> |
| **Physical activity** | 100: ≥10.0 MET hours/week<br>90: 8.0-9.9 MET hours/week<br>80: 6.0-7.9 MET hours/week<br>60: 4.0-5.9 MET hours/week<br>40: 2.0-3.9 MET hours/week<br>20: 0.1-1.9 MET hours/week<br>0: 0 MET hours/week |
| **Diet** | 100: AHEI-2010 score ≥95<sup>th</sup> percentile<br>80: AHEI-2010 score between 75<sup>th</sup>-94<sup>th</sup> percentile<br>50: AHEI-2010 score between 50<sup>th</sup>-74<sup>th</sup> percentile<br>25: AHEI-2010 score between 25<sup>th</sup>-49<sup>th</sup> percentile<br>0: AHEI-2010 score <25<sup>th</sup> percentile |
| **Sleep health** | 100: 7-<9 hours per night<br>90: 9-<10 hours per night<br>70: 6-<7 hours per night<br>40: 5-<6 or ≥10 hours per night<br>20: 4-<5 hours per night<br>0: <4 hours per night |

Abbreviations: AHEI-2010, alternative healthy eating index 2010; BMI, body mass index; CVH, cardiovascular health; DBP, diastolic blood pressure; HbA1c, glycohemoglobin; HDL, high-density lipoprotein; MET, metabolic equivalent of task; SBP, systolic blood pressure.

### Assessment of Overall CVH

The outcome in the study is the overall CVH based on all eight LE8 metrics. We generated both a continuous and two binary measures of overall CVH. The continuous overall CVH score was calculated by averaging scores of all eight LE8 metrics (range: 0 to 100). In addition, we also categorized the continuous CVH score into three categories (i.e., ≥80: high, 50-80: moderate, and <50: low), and two binary outcomes were generated comparing individuals with (1) high CVH vs. moderate or low CVH and (2) low CVH vs. moderate or high CVH.

### Assessment of Predictors

Figure S1 shows the availabilities of each predictor in NHS, NHSII, and HPFS. We first included predictors that are widely available in NHS, NHSII, and HPFS. These predictors included (1) demographic factors such as age (years), sex (female or male), race/ethnicity (non-Hispanic white, non-Hispanic black, Hispanic, and others), (2) CVH-related factors (measured biennially) such as self-reported hypertension (yes or no), self-reported diabetes (yes or no), and self-reported hypercholesterolmia (yes or no), and (3) CVH metrics (measured biennially) including BMI (both the original BMI value and BMI score defined by LE8) and nicotine exposure (defined by LE8). We further included other CVH metrics that are less frequently collected (i.e., approximately every 4 years) in NHS, NHSII, and HPFS as predictors (Figure S1), including self-reported blood pressure, physical activity, diet, and sleep health assessed based on LE8.

### Statistical Analyses

Descriptive analyses were conducted to examine the distribution of participants’ demographics, individual CVH metrics, and overall CVH. Two groups of models were trained separately using data from the cohorts (i.e., NHS, NHSII, and HPFS) and NHANES. Figure 1 shows the model training and testing pipelines. Each group of models contain 16 sets of models each with different predictors: we start by training the base models which included predictors that are routinely collected in NHS, NHSII, and HPFS, such as demographic factors (i.e., age, sex, race/ethnicity), CVH-related factors (i.e., hypertension, hypercholesterolemia, and diabetes), as well as CVH metrics (i.e., BMI and nicotine exposure). We then further included CVH metrics (i.e., blood pressure, physical activity, diet, and sleep health) that are less frequently collected as predictors in additional models (15 sets of models). Of note, percentiles of AHEI-2010 scores were generated separately in the cohorts (i.e., NHS, NHSII, and HPFS) and NHANES for model trainings, and the corresponding cut-points were used to determine diet status in external validations. All models were trained using gradient boosting decision trees implemented by CatBoost (gradient boosting with categorical features support), a highly efficient ensemble-based machine learning model.^61^ Following the best practice in the field, we randomly split the data into a training set (80%) and a testing set (20%). The training sets were used to tune hyperparameters (i.e., number of iterations, number of trees, learning rate, L2 regularization, tree depth, and border count) using grid searches based on 4-fold cross-validated RMSEs (root mean square errors) for the continuous overall CVH score and AUCs (areas under the receiver operator characteristic curve) for the two binary outcomes (i.e., high CVH vs. moderate/low CVH and low CVH vs. moderate/high CVH). The testing set was then used to perform internal validation. External validations were also conducted using external testing data (e.g., models trained using NHS, NHSII, and HPFS data were externally validated using NHANES data and vice versa). To examine the robustness of model performance in different cohorts, we also generated stratified internal validation results in NHS, NHSII, and HPFS, separately.

**Figure 1.**
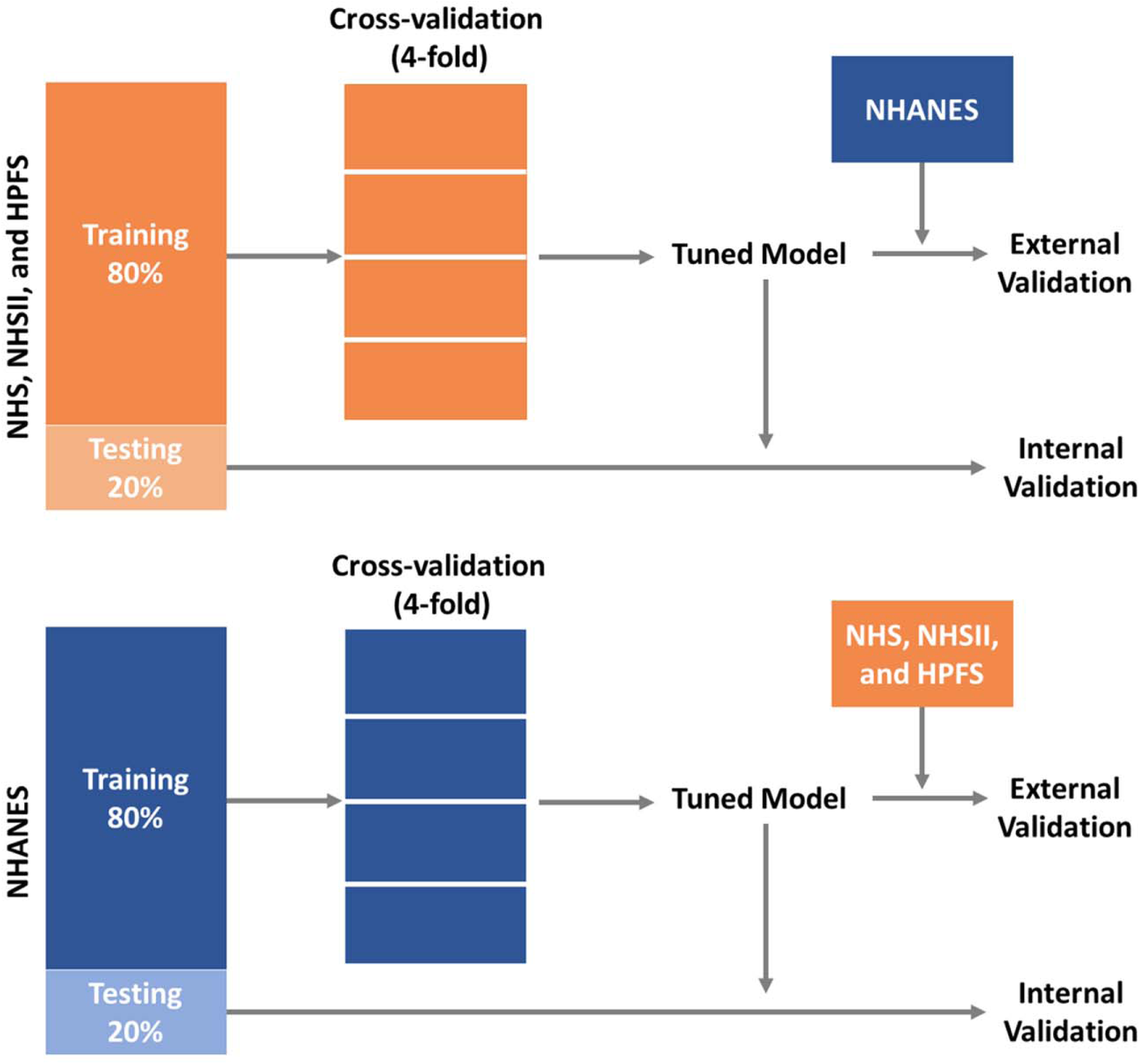
Training and validation pipelines for prediction models of CVH using data from NHS, NHSII, and HPFS, and the 2005-2016 NHANES. Abbreviations: CVH, cardiovascular health; HPFS, Health Professional’s Follow-up Study; NHANES: the National Health and Nutrition Examination Survey; NHS, Nurses’ Health Study; NHSII, Nurses’ Health Study II.

To further examine the performance of this approach in real world settings, we conducted sensitivity analyses by assessing whether the predicted CVH scores can generate consistent effect estimates in associational studies. Cox proportional hazards models were used to assess the associations between all-cause mortality and both the observed and predicted LE8 scores in the internal testing sets in NHS, NHSII, and HPFS as well as the NHANES. Hazard ratios (HR) with 95% confidence intervals (CIs) were generated. To account for the complex survey design of the NHANES, a 12-year weight was calculated by dividing the original two-year weight by 6 for each individual. Models were adjusted for age (continuous), sex (female and male), and race/ethnicity (non-Hispanic White, non-Hispanic Black, Hispanic, and others), and marital status (never married, married or living with partner, and previously married). In addition, in the NHANES, we further adjusted for education (< high school, high school or equivalent, some college, college/graduate or above) and family poverty income ratio (PIR: <1, 1-2, and ≥2).

It has been suggested that the newly introduced LE8 score (0-100 points) is highly correlated with the previous LS7 score (0-14 points).^16^ To assess the robustness of our approach, we have conducted sensitivity analyses using CVH measures based on LS7 as the outcomes (Table S2). Specifically, the seven individual LS7 metrics (i.e., blood pressure, HbA1c, total cholesterol, smoking, BMI, physical activity, and diet) were categorized into 3 levels: poor (0 point), intermediate (1 point), and ideal (2 points). A continuous CVH score was then calculated by summing up scores of all the seven metrics (range: 0 to 14). We also generated seven binary measures of overall CVH based on the number of ideal CVH metrics. We used the same modelling pipeline for LS7, with a total of 8 sets of predictors. Similarly, Cox proportional hazards models were also fitted in the internal testing sets to assess the associations between all-cause mortality and both the observed and predicted LS7 scores.

**Table 2.** Characteristics of participants in NHS, NHSII, and HPFS, and the 2005-2016 NHANES included in developing prediction models of Life’s Essential 8 score.

| Characteristics |  | NHS, NHSII, and HPFS Cohorts |  |  |  | NHANES<br>(n=27,194) |
| --- | --- | --- | --- | --- | --- | --- |
|  |  | NHS<br>(n=4,114) | NHSII<br>(n=676) | HPFS<br>(n=798) | Total<br>(n=5,588) |  |
|  |  | Mean ± SD / n (%) |  |  |  |  |
| Age (years) |  | 59.6 ± 6.5 | 45.2 ± 4.1 | 62.9 ± 8.7 | 58.3 ± 8.3 | 48.8 ± 17.8 |
| BMI (kg/m<sup>2</sup>) |  | 26.1 ± 5.1 | 26.0 ± 5.7 | 25.9 ± 3.3 | 26.1 ± 5.0 | 29.0 ± 6.8 |
| Sex |  |  |  |  |  |  |
|  | Male | 0 (0.0) | 0 (0.0) | 798 (100.0) | 798 (14.3) | 13,219 (48.6) |
|  | Female | 4,114 (100.0) | 676 (100.0) | 0 (0.0) | 4,790 (85.7) | 13,975 (51.4) |
| Race/ethnicity |  |  |  |  |  |  |
|  | Non-Hispanic White | 3,872 (94.1) | 656 (97.0) | 426 (53.4) | 4,954 (88.7) | 12,180 (44.8) |
|  | Non-Hispanic Black | 19 (0.5) | 5 (0.7) | 0 (0.0) | 24 (0.4) | 5,471 (20.1) |
|  | Hispanic | 30 (0.7) | 7 (1.0) | 4 (0.5) | 41 (0.7) | 7,059 (26.0) |
|  | Others | 193 (4.7) | 8 (1.2) | 368 (46.1) | 569 (10.2) | 2,484 (9.1) |
| Hypertension |  |  |  |  |  |  |
|  | No | 3,041 (73.9) | 605 (89.5) | 602 (75.4) | 4,248 (76.0) | 17,721 (65.2) |
|  | Yes | 1,073 (26.1) | 71 (10.5) | 196 (24.6) | 1,340 (24.0) | 9,437 (34.7) |
|  | Missing | 0 (0) | 0 (0) | 0 (0) | 0 (0) | 36 (0.1) |
| Diabetes |  |  |  |  |  |  |
|  | No | 3,574 (86.9) | 658 (97.3) | 760 (95.2) | 4,992 (89.3) | 23,216 (85.4) |
|  | Yes | 540 (13.1) | 18 (2.7) | 38 (4.8) | 596 (10.7) | 3,978 (14.6) |
| Hypercholesterolemia |  |  |  |  |  |  |
|  | No | 2,536 (61.6) | 577 (85.4) | 570 (71.4) | 3,683 (65.9) | 14,010 (51.5) |
|  | Yes | 1,578 (38.4) | 99 (14.6) | 228 (28.6) | 1,905 (34.1) | 8,874 (32.6) |
|  | Missing | 0 (0) | 0 (0) | 0 (0) | 0 (0) | 4,310 (15.8) |
| Overall CVH |  |  |  |  |  |  |
|  | *LE8 score (0-100)* | 65.4 ± 13.1 | 73.8 ± 14.2 | 60.0 ± 10.3 | 65.6 ± 13.4 | 61.6 ± 14.3 |
|  | *Categorical measure* |  |  |  |  |  |
|  | Low (LE8 score<50) | 503 (12.2) | 37 (5.5) | 134 (16.8) | 674 (12.1) | 5,743 (21.1) |
|  | Moderate (LE8 score 50-80) | 3,040 (73.9) | 379 (56.1) | 653 (81.8) | 4,072 (72.9) | 18,424 (67.8) |
|  | High (LE8 score≥80) | 571 (13.9) | 260 (38.5) | 11 (1.4) | 842 (15.1) | 3,027 (11.1) |
| Individual LE8 metric scores |  |  |  |  |  |  |
|  | *Blood pressure* | 45.9 ± 28.9 | 65.1 ± 29.8 | 45.3 ± 24.4 | 48.1 ± 29.1 | 59.6 ± 31.8 |
|  | *HbA1c* | 80.9 ± 28.0 | 95.7 ± 15.8 | 1.5 ± 0.6 | 71.4 ± 38.0 | 80.4 ± 27.1 |
|  | *Blood lipids* | 42.1 ± 33.3 | 63.1 ± 34.3 | 53.7 ± 31.8 | 46.3 ± 34.0 | 64.4 ± 31.0 |
|  | *Nicotine exposure* | 71.0 ± 35.3 | 81.2 ± 32.0 | 79.7 ± 26.5 | 73.4 ± 34.0 | 70.5 ± 39.5 |
|  | *BMI* | 75.8 ± 29.1 | 77.3 ± 30.1 | 78.2 ± 22.5 | 76.3 ± 28.4 | 60.5 ± 33.5 |
|  | *Physical activity* | 79.5 ± 28.9 | 80.7 ± 29.6 | 89.6 ± 23.8 | 81.1 ± 28.5 | 44.7 ± 46.7 |
|  | *Diet* | 39.4 ± 30.8 | 39.0 ± 32.5 | 42.0 ± 33.1 | 39.8 ± 31.3 | 31.9 ± 26.6 |
|  | *Sleep health* | 88.5 ± 19.5 | 88.5 ± 21.8 | 89.7 ± 20.2 | 88.6 ± 19.9 | 80.8 ± 25.8 |

Abbreviations: BMI, body mass index; CVH, cardiovascular health; HbA1c, glycohemoglobin, LE8, Life's Essential 8.

All analyses were conducted using R version 4.1.0 with CatBoost models implemented using the “catboost” R package.^62^

## Results

A total of 5,588 and 27,194 participants from the NHS, NHSII, and HPFS cohorts and the 2005-2016 NHANES with complete information on all eight CVH metrics were included in this study, respectively. Table 2 shows the distributions of participants’ demographic characteristics, medical history, overall LE8 score, and individual LE8 metric scores. Compared with participants in the NHANES, participants in NHS, NHSII, and HPFS were older, more likely to be non-Hispanic White, less likely to have hypertension, diabetes, and hypercholesterolemia, and more likely to have better overall CVH. In addition, participants in NHS, NHSII, and HPFS were also more likely to have more optimal individual CVH metrics including BMI, nicotine exposure, physical activity, diet, and sleep health, while those in the NHANES were more likely to have better status in blood pressure, HbA1c, and blood lipids (all p<0.001).

Hyperparameters tuned based on grid searches are presented in Tables S3 and S4 for the models trained using the cohorts and NHANES, respectively. Figure 2 shows the performance of models to estimate the continuous overall CVH score based on LE8. Internally and externally validated RMSEs of 8.06 and 16.72 were observed, respectively, in base models trained using the cohorts. Similarly, in base models trained using NHANES, internally and externally validated RMSEs of 9.21 and 18.33 were observed. Models additionally including physical activity, diet, blood pressure, and sleep health had the best internally validated RMSEs (3.94 in the best model trained using the cohorts, and 4.24 in the best model trained using NHANES). Models trained using the cohorts with additional predictors including blood pressure and sleep health had the best externally validated RMSE of 14.25, while models trained using NHANES had best externally validated RMSE of 10.39 with additional predictors including physical activity, diet, blood pressure, and sleep health.

**Figure 2.**
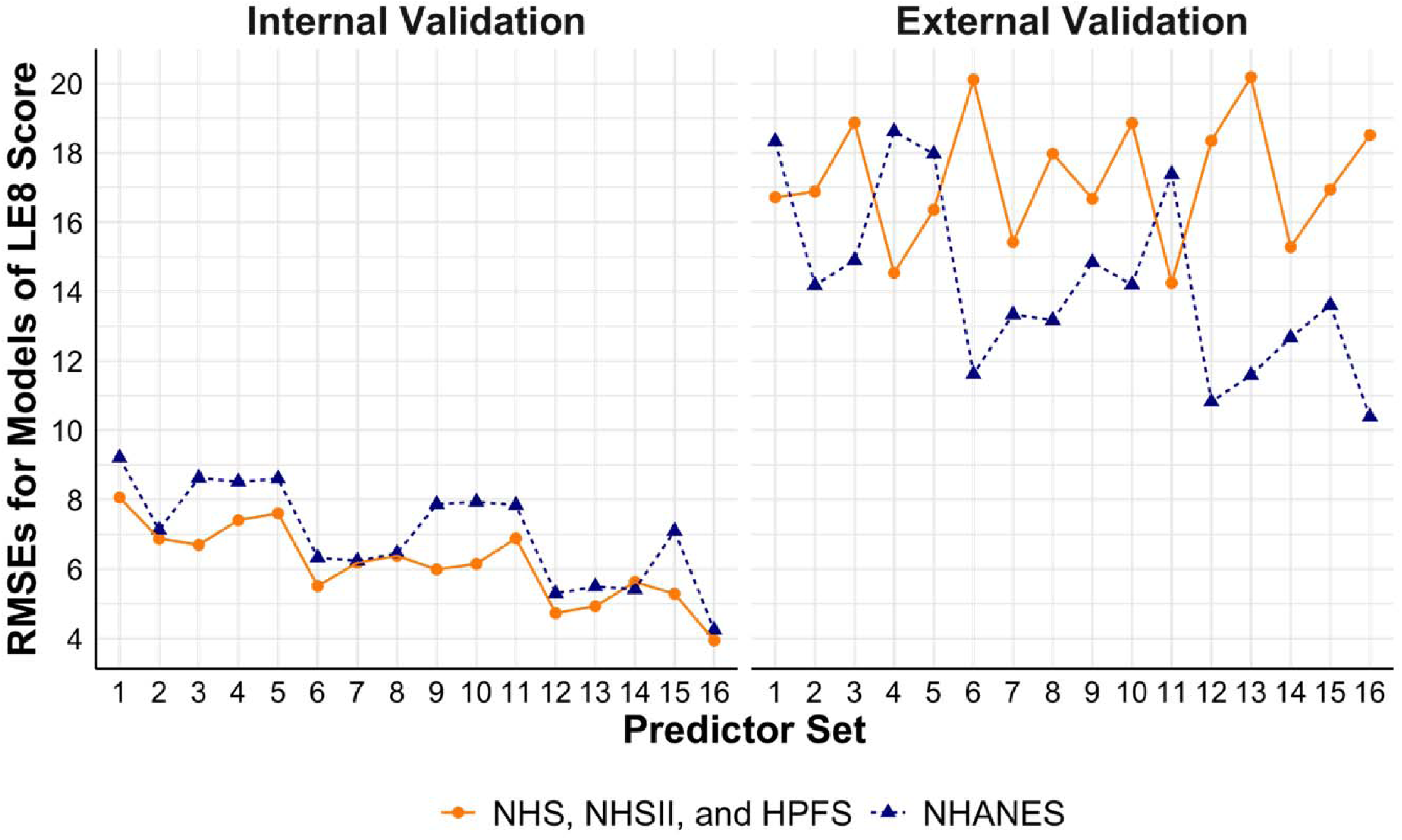
Performance of models to estimate continuous LE8 score using NHS, NHSII, and HPFS (n=5,588), and NHANES (n=27,194). Set 1 (i.e., base model): Age, sex, race/ethnicity, BMI, smoking, hypertension, hypercholesterolemia, and diabetes; Set 2: + physical activity; Set 3: + diet; Set 4: + blood pressure; Set 5: + sleep health; Set 6: + physical activity + diet; Set 7: + physical activity + blood pressure; Set 8: + physical activity + sleep health; Set 9: + diet + blood pressure; Set 10: + diet + sleep health; Set 11: + blood pressure + sleep health; Set 12: + physical activity + diet + blood pressure; Set 13: + physical activity + diet + sleep health; Set 14: + physical activity + blood pressure + sleep health; Set 15: + diet + blood pressure + sleep health; Set 16: + physical activity + diet + blood pressure + sleep health. Abbreviations: BMI, body mass index; CVH, cardiovascular health; HPFS, Health Professional’s Follow-up Study; LE8, Life’s Essential 8; NHANES: the National Health and Nutrition Examination Survey; NHS, Nurses’ Health Study; NHSII, Nurses’ Health Study II; RMSE, root mean square error.

Figures 3 shows the performance of models to estimate binary CVH outcomes. In models trained using the cohorts, the base models had validated AUCs of 0.91 and 0.92 (internal) and 0.56 and 0.60 (external) for high vs. moderate/low CVH and low vs. moderate/high CVH, respectively. Similarly, the base models trained using NHANES had internally validated AUCs of 0.91 and 0.89 and externally validated AUCs of 0.70 and 0.51 for the two binary CVH outcomes, respectively. Models with additional predictors such as physical activity, diet, blood pressure, and sleep health had better performance, with the best validated AUCs of 0.98 and 0.98 (internal) and 0.89 and 0.78 (external) in models trained using the cohorts, and 0.99 and 0.97 (internal) and 0.89 and 0.77 (external) in models trained using NHANES for the two binary CVH outcomes, respectively.

**Figure 3.**
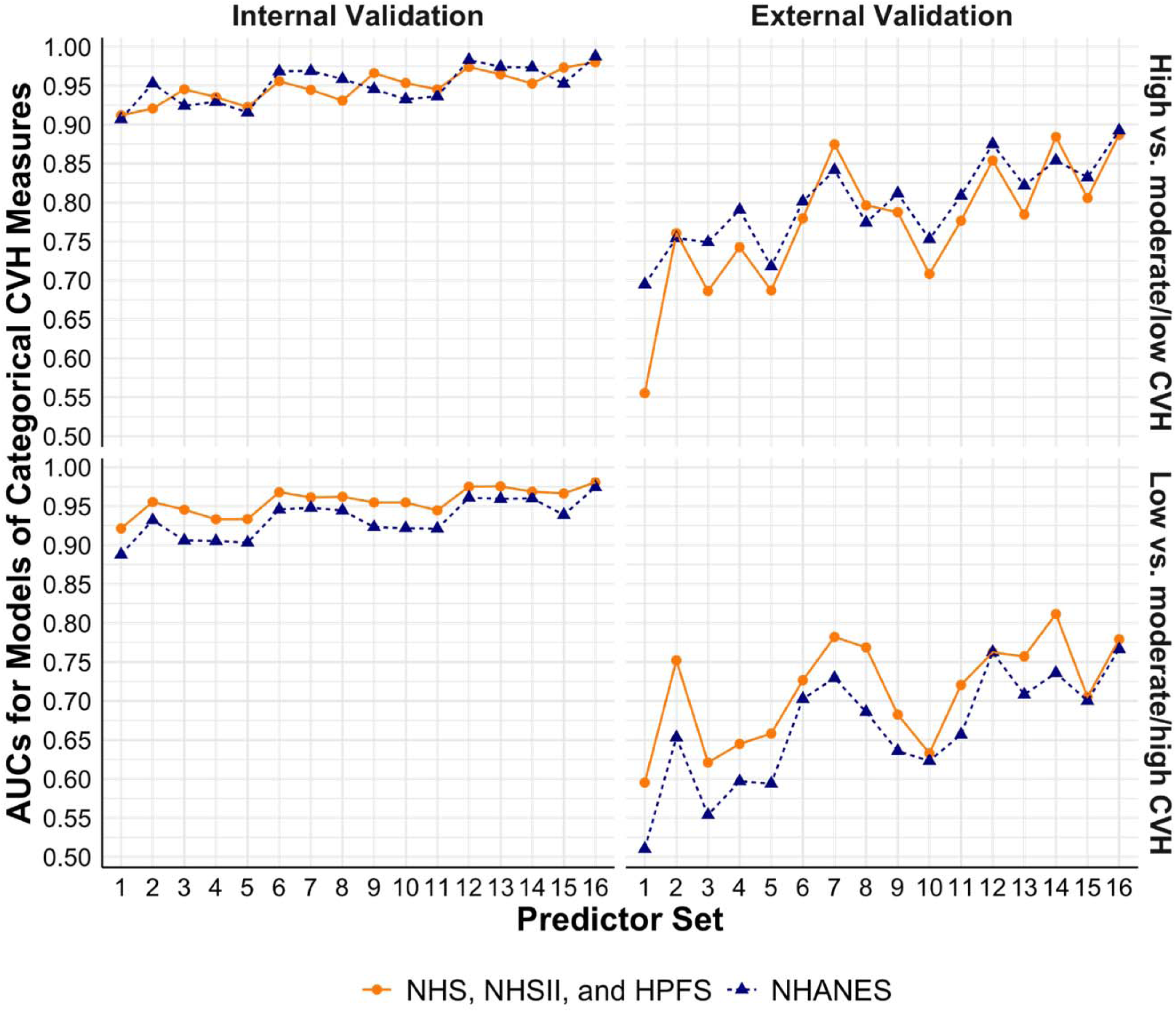
Performance of models to estimate categorical CVH measures based on LE8 score using NHS, NHSII, and HPFS (n=5,588), and NHANES (n=27,194). Set 1 (i.e., base model): Age, sex, race/ethnicity, BMI, smoking, hypertension, hypercholesterolemia, and diabetes; Set 2: + physical activity; Set 3: + diet; Set 4: + blood pressure; Set 5: + sleep health; Set 6: + physical activity + diet; Set 7: + physical activity + blood pressure; Set 8: + physical activity + sleep health; Set 9: + diet + blood pressure; Set 10: + diet + sleep health; Set 11: + blood pressure + sleep health; Set 12: + physical activity + diet + blood pressure; Set 13: + physical activity + diet + sleep health; Set 14: + physical activity + blood pressure + sleep health; Set 15: + diet + blood pressure + sleep health; Set 16: + physical activity + diet + blood pressure + sleep health. Abbreviations: AUC, area under the receiver operator characteristic curve; BMI, body mass index; CVH, cardiovascular health; HPFS, Health Professional’s Follow-up Study; LE8, Life’s Essential 8; NHANES: the National Health and Nutrition Examination Survey; NHS, Nurses’ Health Study; NHSII, Nurses’ Health Study II.

Tables S5 and S6 show the detailed results for each model. Consistent results were observed in internal validations by cohort (Table S7).

Figure 4 presents the HRs and 95% CIs for all-cause mortality. In the cohorts, one unit increase in the observed LE8 score was associated with significantly lower hazards of all-cause mortality (HR: 0.982, 95% CI: 0.976-0.989). Consistent results were observed in models using predicted LE8 scores based on different sets of predictors. Similarly, in the NHANES, no statistically significant difference was found between the associations of all-cause mortality with the observed and predicted LE8 scores.

**Figure 4.**
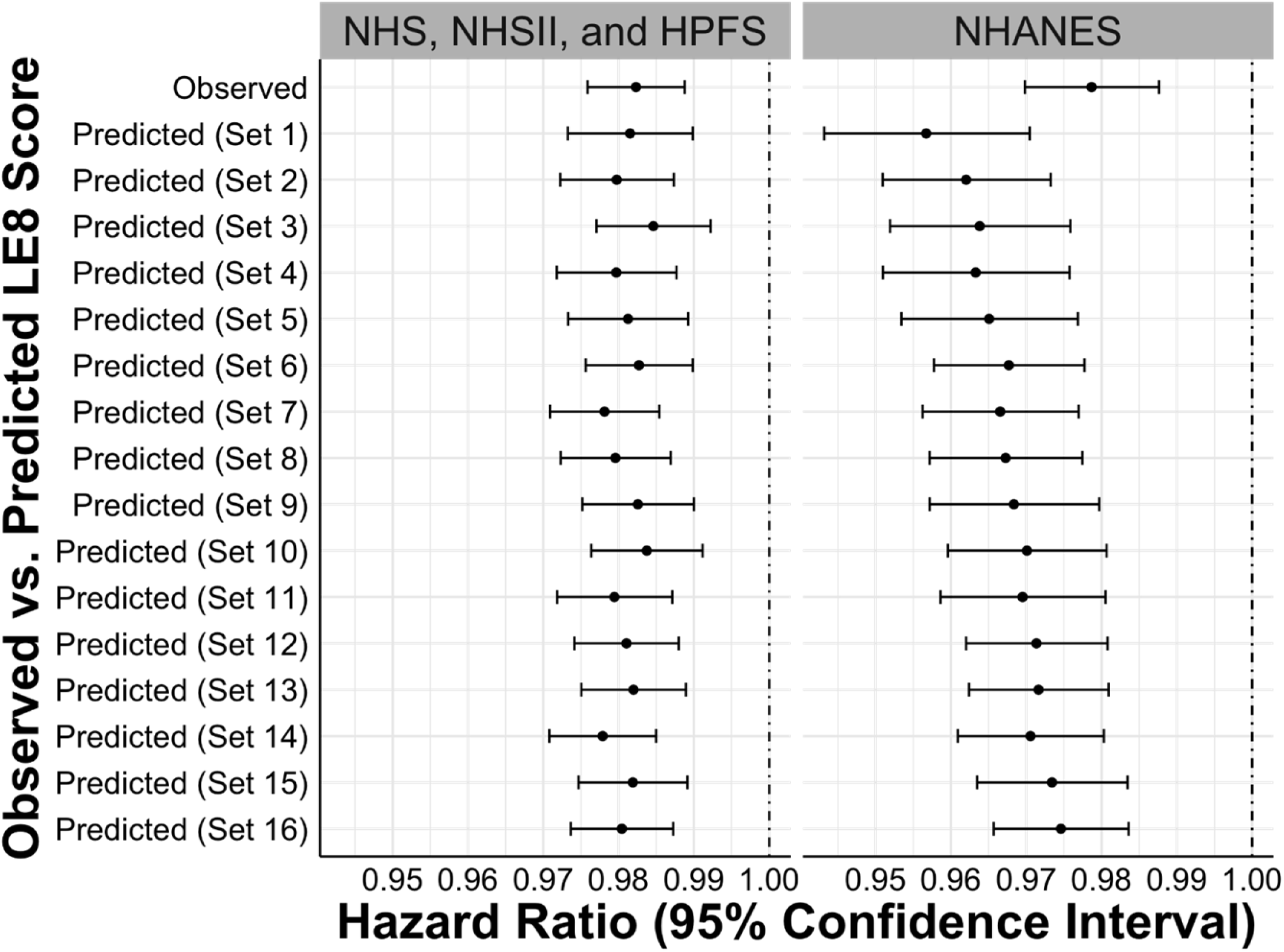
Hazard ratios and 95% confidence intervals for the associations between observed vs. predicted LE8 scores and all-cause mortality in internal testing sets of NHS, NHSII, and HPFS (n=5,588), and NHANES (n=27,194). Set 1 (i.e., base model): Age, sex, race/ethnicity, BMI, smoking, hypertension, hypercholesterolemia, and diabetes; Set 2: + physical activity; Set 3: + diet; Set 4: + blood pressure; Set 5: + sleep health; Set 6: + physical activity + diet; Set 7: + physical activity + blood pressure; Set 8: + physical activity + sleep health; Set 9: + diet + blood pressure; Set 10: + diet + sleep health; Set 11: + blood pressure + sleep health; Set 12: + physical activity + diet + blood pressure; Set 13: + physical activity + diet + sleep health; Set 14: + physical activity + blood pressure + sleep health; Set 15: + diet + blood pressure + sleep health; Set 16: + physical activity + diet + blood pressure + sleep health. Abbreviations: BMI, body mass index; CVH, cardiovascular health; HPFS, Health Professional’s Follow-up Study; LE8, Life’s Essential 8; NHANES: the National Health and Nutrition Examination Survey; NHS, Nurses’ Health Study; NHSII, Nurses’ Health Study II.

To further assess the robustness of our approach, we conducted sensitivity analyses using CVH measures based on LS7 (Table S2). Table S8 shows the distributions of demographic characteristics, medical history, overall LS7 CVH, and individual LS7 metrics. Data from 1999-2004 NHANES were not used in the main analyses based on LE8 since sleep health was not available, however, they were included in the sensitivity analyses. A total of 8,500 and 39,933 participants from the cohorts and the 1999-2016 NHANES with complete information on all seven LS7 metrics were included in this study, respectively. Consistent with findings for LE8, participants in the cohorts were less likely to have hypertension, diabetes, and hypercholesterolemia and had better overall CVH, compared with participants in the NHANES. Participants in the cohorts were also more likely to have ideal status for individual CVH metrics including BMI, cigarette smoking, physical activity, and diet, while those in the NHANES were more likely to have ideal blood pressure and total cholesterol (all p<0.001).

Tables S9 and S10 show tuned hyperparameters for the models trained using the cohorts and NHANES, respectively. Figures S2 and S3 show the performance of models for the continuous CVH score and binary overall CVH measures assessed by LS7. In base models trained using the cohorts, validated RMSEs of 1.47 (internal) and 2.37 (external) and validated AUCs ranging from 0.85 to 0.98 (internal) and 0.74 to 0.90 (external) were observed. Similarly, the base models trained using NHANES had validated RMSEs of 1.55 (internal) and 3.19 (external) and validated AUCs ranging from 0.85 to 0.97 (internal) and 0.77 to 0.87 (external). Models with additional predictors such as physical activity, diet, and/or blood pressure had better performance, with the best validated RMSEs of 0.86 (internal) and 1.81 (external) and validated AUCs ranging from 0.96 to 0.99 (internal) and 0.79 to 0.94 (external) in models trained using the cohorts, and the best validated RMSEs of 0.82 (internal) and 1.92 (external) and validated AUCs ranging from 0.95 to 0.99 (internal) and 0.89 to 0.98 (external) in models trained using NHANES. Tables S11 and S12 shows the detailed results for each model. Results of stratified internal validations for models of LS7 in each of the cohorts are shown in Table S13.

Figure S4 presents associations between all-cause mortality and the observed and predicted LS7 scores. Similar to the results observed for the LE8 scores, no statistically significant difference was observed in the associations based on the observed vs. predicted LE7 scores.

## Discussion

Leveraging data from three nationwide prospective cohorts (i.e., NHS, NHSII, and HPFS) and a series of cross-sectional nationally representative data from the NHANES, we developed and validated several sets of models to estimate individuals’ overall CVH status defined by LE8 when not all eight metrics are available. We found that information routinely collected and widely available in many research studies and clinical settings (e.g., age, sex, race/ethnicity, BMI, nicotine exposure, hypertension, hypercholesterolemia, and diabetes) can be used to accurately estimate individuals’ overall CVH status. Consistent results were observed in sensitivity analyses defining CVH outcomes based on LS7. In addition, the predicted CVH scores can generate consistent effect estimates in associational studies as the observed CVH scores.

Both the original LS7 and the recently updated LE8 metrics introduced by the AHA emphasize primordial prevention, and have great potential to guide and improve CVD prevention.^3,5^ It has been shown that individuals’ overall CVH declines with age.^14–16^ A recent pooled cohort analysis on trajectories of clinical CVH scores (based on BMI, blood pressure, cholesterol, and blood glucose) identified two inflection points in late adolescence (i.e., 16.9 years) and early middle age (i.e., 37.2 years) during which the decline of CVH accelerates.^28^ It is thus important to identify and understand factors contributing to CVH declines at different stages of life. However, due to the challenges to simultaneously measure all eight LE8 (or seven LS7) CVH metrics over time, most existing studies on CVH are cross-sectional,^14,17–22^ and the few longitudinal studies which examined individuals’ CVH trajectories over time either only had CVH sparsely measured over time (e.g., ≤3 time points in ≥10 years) or used modified versions of LE8 or LS7 (e.g., the clinical CVH score).^23–28^ The models developed and validated in this study provide a cost-effective and feasible solution to enable longitudinal assessment of CVH trajectories in multiple settings when not all eight LE8 (or seven LS7) CVH metrics are available.

In this study, we observed great model performance in internal validations for different predictors-outcome pairs in models either trained using the cohorts (i.e., NHS, NHSII, and HPFS) or the NHANES. This is not unexpected as many of the CVH metrics included in LE8 and LS7 are highly correlated, and therefore, it is plausible to use some but not all eight LE8 (or seven LS7) metrics along with other CVH-related factors to estimate individuals’ overall CVH. This is supported by results from a recent study, which used 13-year electronic health records with measures of five CVH metrics (i.e., smoking, BMI, blood pressure, glucose, and cholesterol) and found that future individual CVH metrics can be reliably predicted using previous measures of these metrics.^27^ In addition, we also showed that the predicted CVH scores can generate consistent effect estimates in associational studies as the observed CVH scores. Our findings suggest that in research and clinical settings without all eight LE8 (or seven LS7) CVH metrics measured at every time point, using the few CVH metrics and related factors routinely collected can accurately estimate individuals’ overall CVH, making it feasible to examine trajectories of overall CVH over time.

Compared with the results from internal validations, the models performed relatively worse in external validations, which may be mainly caused by differences between the data used in internal and external validations, including (1) different study populations (e.g., NHS, NHSII, and HPFS included only health professions and participants are older, while the NHANES included the general population), and (2) different measurement methods of individual CVH metrics and predictors (e.g., blood pressures were based on self-report in NHS, NHSII, and HPFS, while the NHANES used the average blood pressure from consecutive measurements). These results suggest that while directly using off-the-shelf models pretrained using other data sources (e.g., NHANES) are feasible, when possible, it is ideal to retrain and validate models for specific research or clinical settings, especially when the targeted populations or measurement methods are different from the original data source used to develop the pretrained models. There are several strengths and some limitations to note. This is the first effort to estimate individuals’ overall CVH when not all eight LE8 (or seven LS7) CVH metrics are available. We showed that the few CVH metrics and related factors routinely collected in many research and clinical settings can be used to accurately estimate individuals’ overall CVH. This is especially valuable to longitudinal studies focusing on CVH trajectories as it enables inclusions of data from more time points to better characterize longitudinal changes in overall CVH. It is also clinically relevant by providing a cost-effective and feasible way to track individuals’ CVH over time. In addition, using three large nationwide prospective cohorts (NHS, NHSII, and HPFS) and the nationally representative NHANES, the results observed, and implications drawn from this study are generalizable to other populations and study settings. One limitation to note is the relatively worse model performance in external validations, which suggested that directly applying off-the-shelf models pretrained using data from other population or setting may yield less accurate estimations. However, the consistently great model performance observed in internal validations using both the cohorts (i.e., NHS, NHSII, and HPFS) and NHANES data provide strong evidence suggesting that individuals’ overall CVH can be accurately estimated with retrained and fine-tuned models for specific research or clinical settings.

## Conclusions

Using data from three large nationwide prospective cohorts (i.e., NHS, NHSII, and HPFS) and a nationally representative survey (i.e., NHANES), we showed that CVH-related factors routinely measured in many research and clinical settings can be used to accurately estimate individuals’ overall CVH even when not all eight LE8 (or seven LS7) metrics are available. In summary, the approach introduced in this study provides a cost-effective and feasible way to estimate individuals’ overall CVH in multiple settings and is especially valuable to characterize individuals’ CVH trajectories over time.

## Data Availability

NHANES data are publicly available. We provide access to the data and resources of the Nurses Health Studies (NHS, NHSII) and Health Professionals Follow-Up Study (HPFS) for use by external investigators and consortia to optimize scientific inquiry while preserving participant confidentiality with guidance and approval from our Institutional Review Board. As an overview, external investigators access NHS, NHSII, and HPFS data in one of three ways: 1) through the NHS, NHSII, and HPFS Data Repository, the external investigator is granted a login to our computer system, accesses and analyzes our data, 2) the external investigator collaborates directly with an internal investigator and programmer who conduct the analyses on the external investigator's behalf, or 3) a specific, limited dataset is created to send to the external collaborator. Given the greater flexibility provided by the 1st option with direct access to all cohort data, the 2nd and 3rd options are not utilized as frequently, except for consortia projects pooling data.

https://www.hsph.harvard.edu/hpfs/

http://nurseshealthstudy.org/

https://www.cdc.gov/nchs/nhanes/index.htm

## Non-standard Abbreviations and Acronyms

ACC: American College of Cardiology
AHEI-2010: Alternative healthy eating index 2010\
CI: confidence interval
CVD: Cardiovascular disease
CVH: Cardiovascular health
HPFS: Health Professionals Follow-up Study
HR: hazard ratio
LE8: Life’s Essential 8
LS7: Life’s Simple 7
MET: Metabolic equivalent of task
NHANES: National Health and Nutrition Examination Survey
NHS: Nurses’ Health Study
NHSII: Nurses’ Health Study II

---

**Table S1.** Timing of blood sample collections and questionnaires used to assess CVH metrics in NHS, NHSII, and HPFS.

| Cohort | Blood Sample<br>(Year of<br>Collection) | Questionnaire from Closest Follow-up Cycles with Available Data (Year of Collection) |  |  |  |  |  |  |
| --- | --- | --- | --- | --- | --- | --- | --- | --- |
|  |  | Blood<br>Pressure | BMI | Cigarette<br>Smoking | Physical<br>Activity | Diet | Sleep | Medications |
| NHS | 1989-1991 | 1990 | 1990 | 1990 | 1992 | 1990 | 1986 | 1988 |
| NHSII | 1996-1999 | 1999 | 1999 | 1999 | 2001 | 1999 | 2001 | 2001 |
| HPFS | 1993-1995 | 1992, 1996 | 1994, 1996 | 1994, 1996 | 1994, 1996 | 1994 | 2000 | 1994, 1996 |

Abbreviations: BMI, body mass index; CVH, cardiovascular health; HPFS, Health Professionals Follow-up Study; NHS, Nurses' Health Study; NHSII, Nurses' Health Study II.

**Table S2.** Definitions of poor, intermediate, and ideal CVH metrics based on Life’s Simple 7.

| **CVH metrics** | **Poor** | **Intermediate** | **Ideal** |
| --- | --- | --- | --- |
| Blood pressure | SBP≥135mmHg or<br>DBP≥85mmHg | SBP 115-134mmHg, or DBP 75-84mmHg, or<br>treated to goal | SBP<115mmHg and DBP<75mmHg,<br>untreated |
| HbA1c | >6.4% | 5.7-6.4% or treated to goal | <5.7%, untreated |
| Total cholesterol | ≥240mg/dL | 200-239mg/dL or treated to goal | <200mg/dL, untreated |
| Smoking | Current smoking | Former, quit ≤12 months previously | Never or quit >12 months previously |
| BMI | ≥30.0kg/m<sup>2</sup> | 25.0-29.9kg/m<sup>2</sup> | <25.0kg/m<sup>2</sup> |
| Physical activity | None | <10 MET hours/week | ≥10 MET hours/week |
| Diet | AHEI-2010 Tertile 1 | AHEI-2010 Tertile 2 | AHEI-2010 Tertile 3 |

Abbreviations: AHEI-2010, alternative healthy eating index 2010; BMI, body mass index; CVH, cardiovascular health; DBP, diastolic blood pressure; HbA1c, glycohemoglobin; MET, metabolic equivalent of task; SBP, systolic blood pressure.

**Table S3.**
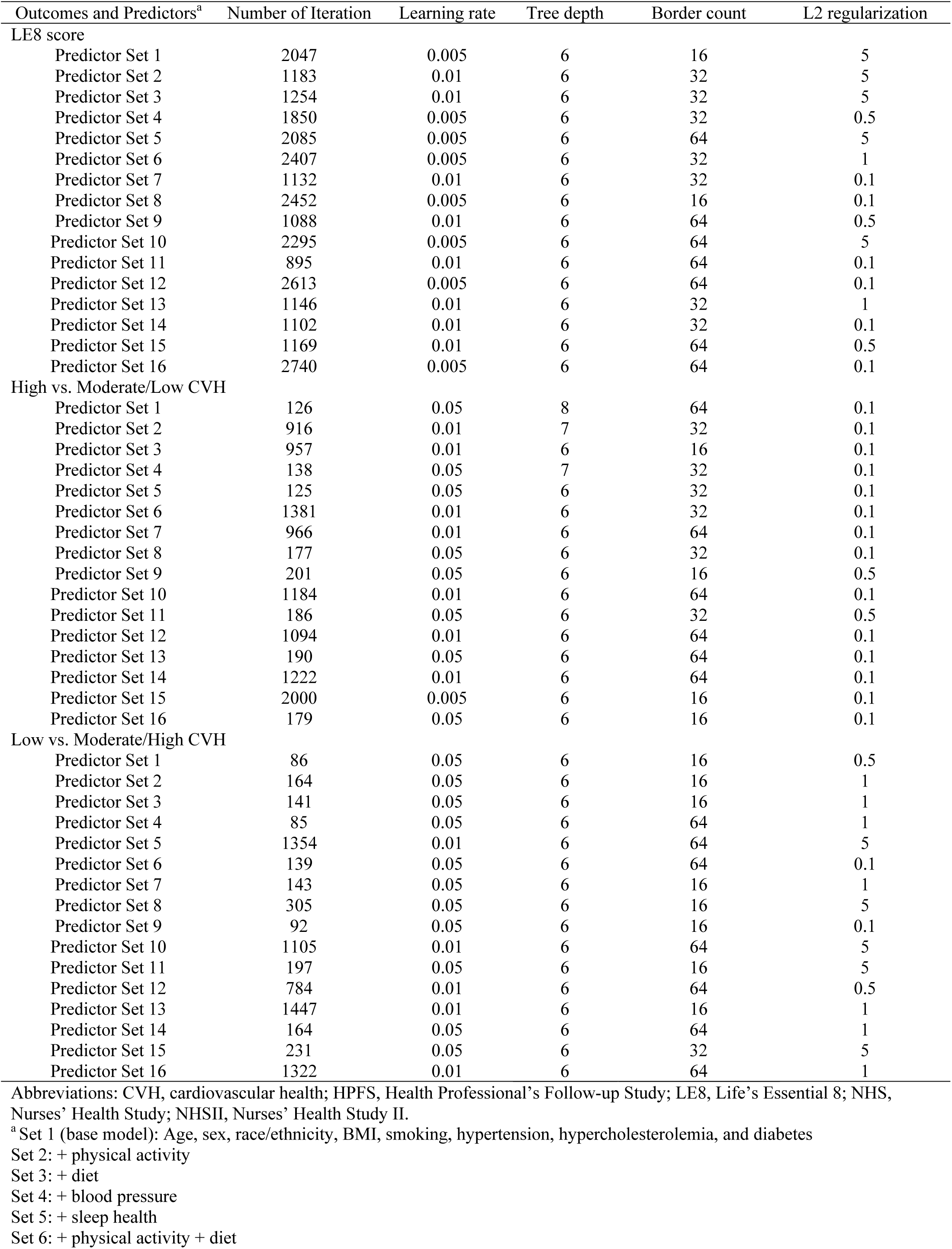
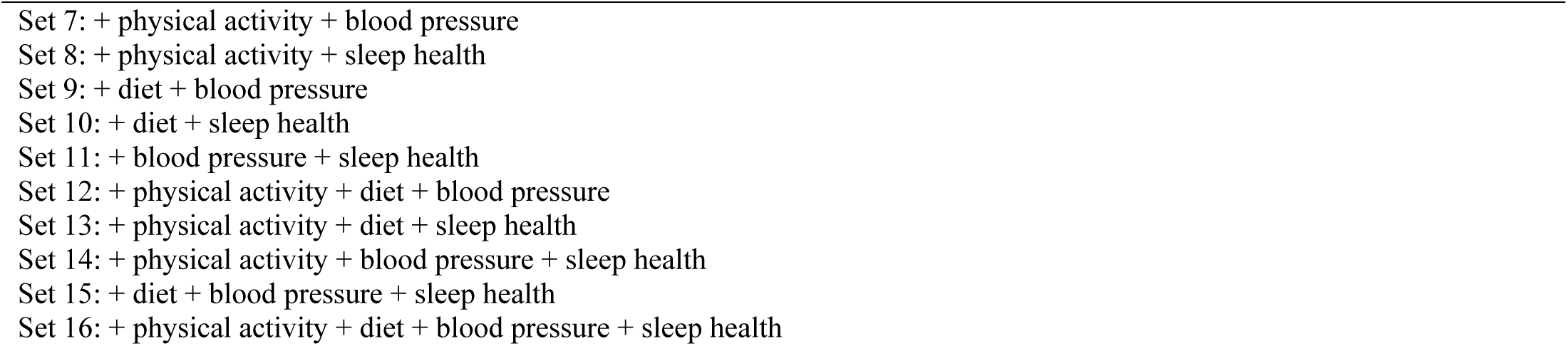
Optimal hyperparameters of predictive models of CVH based on LE8 tuned by cross-validation in the training set using NHS, NHSII, and HPFS (n=5,588).

| Outcomes and Predictors<sup>a</sup> | Number of Iteration | Learning rate | Tree depth | Border count | L2 regularization |
| --- | --- | --- | --- | --- | --- |
| LE8 score |  |  |  |  |  |
| Predictor Set 1 | 2047 | 0.005 | 6 | 16 | 5 |
| Predictor Set 2 | 1183 | 0.01 | 6 | 32 | 5 |
| Predictor Set 3 | 1254 | 0.01 | 6 | 32 | 5 |
| Predictor Set 4 | 1850 | 0.005 | 6 | 32 | 0.5 |
| Predictor Set 5 | 2085 | 0.005 | 6 | 64 | 5 |
| Predictor Set 6 | 2407 | 0.005 | 6 | 32 | 1 |
| Predictor Set 7 | 1132 | 0.01 | 6 | 32 | 0.1 |
| Predictor Set 8 | 2452 | 0.005 | 6 | 16 | 0.1 |
| Predictor Set 9 | 1088 | 0.01 | 6 | 64 | 0.5 |
| Predictor Set 10 | 2295 | 0.005 | 6 | 64 | 5 |
| Predictor Set 11 | 895 | 0.01 | 6 | 64 | 0.1 |
| Predictor Set 12 | 2613 | 0.005 | 6 | 64 | 0.1 |
| Predictor Set 13 | 1146 | 0.01 | 6 | 32 | 1 |
| Predictor Set 14 | 1102 | 0.01 | 6 | 32 | 0.1 |
| Predictor Set 15 | 1169 | 0.01 | 6 | 64 | 0.5 |
| Predictor Set 16 | 2740 | 0.005 | 6 | 64 | 0.1 |
| High vs. Moderate/Low CVH |  |  |  |  |  |
| Predictor Set 1 | 126 | 0.05 | 8 | 64 | 0.1 |
| Predictor Set 2 | 916 | 0.01 | 7 | 32 | 0.1 |
| Predictor Set 3 | 957 | 0.01 | 6 | 16 | 0.1 |
| Predictor Set 4 | 138 | 0.05 | 7 | 32 | 0.1 |
| Predictor Set 5 | 125 | 0.05 | 6 | 32 | 0.1 |
| Predictor Set 6 | 1381 | 0.01 | 6 | 32 | 0.1 |
| Predictor Set 7 | 966 | 0.01 | 6 | 64 | 0.1 |
| Predictor Set 8 | 177 | 0.05 | 6 | 32 | 0.1 |
| Predictor Set 9 | 201 | 0.05 | 6 | 16 | 0.5 |
| Predictor Set 10 | 1184 | 0.01 | 6 | 64 | 0.1 |
| Predictor Set 11 | 186 | 0.05 | 6 | 32 | 0.5 |
| Predictor Set 12 | 1094 | 0.01 | 6 | 64 | 0.1 |
| Predictor Set 13 | 190 | 0.05 | 6 | 64 | 0.1 |
| Predictor Set 14 | 1222 | 0.01 | 6 | 64 | 0.1 |
| Predictor Set 15 | 2000 | 0.005 | 6 | 16 | 0.1 |
| Predictor Set 16 | 179 | 0.05 | 6 | 16 | 0.1 |
| Low vs. Moderate/High CVH |  |  |  |  |  |
| Predictor Set 1 | 86 | 0.05 | 6 | 16 | 0.5 |
| Predictor Set 2 | 164 | 0.05 | 6 | 16 | 1 |
| Predictor Set 3 | 141 | 0.05 | 6 | 16 | 1 |
| Predictor Set 4 | 85 | 0.05 | 6 | 64 | 1 |
| Predictor Set 5 | 1354 | 0.01 | 6 | 64 | 5 |
| Predictor Set 6 | 139 | 0.05 | 6 | 64 | 0.1 |
| Predictor Set 7 | 143 | 0.05 | 6 | 16 | 1 |
| Predictor Set 8 | 305 | 0.05 | 6 | 16 | 5 |
| Predictor Set 9 | 92 | 0.05 | 6 | 16 | 0.1 |
| Predictor Set 10 | 1105 | 0.01 | 6 | 64 | 5 |
| Predictor Set 11 | 197 | 0.05 | 6 | 16 | 5 |
| Predictor Set 12 | 784 | 0.01 | 6 | 64 | 0.5 |
| Predictor Set 13 | 1447 | 0.01 | 6 | 16 | 1 |
| Predictor Set 14 | 164 | 0.05 | 6 | 64 | 1 |
| Predictor Set 15 | 231 | 0.05 | 6 | 32 | 5 |
| Predictor Set 16 | 1322 | 0.01 | 6 | 64 | 1 |

Abbreviations: CVH, cardiovascular health; HPFS, Health Professional's Follow-up Study; LE8, Life's Essential 8; NHS, Nurses' Health Study; NHSII, Nurses' Health Study II.
<sup>a</sup> Set 1 (base model): Age, sex, race/ethnicity, BMI, smoking, hypertension, hypercholesterolemia, and diabetes
Set 2: + physical activity
Set 3: + diet
Set 4: + blood pressure
Set 5: + sleep health
Set 6: + physical activity + diet

**Table S4.**
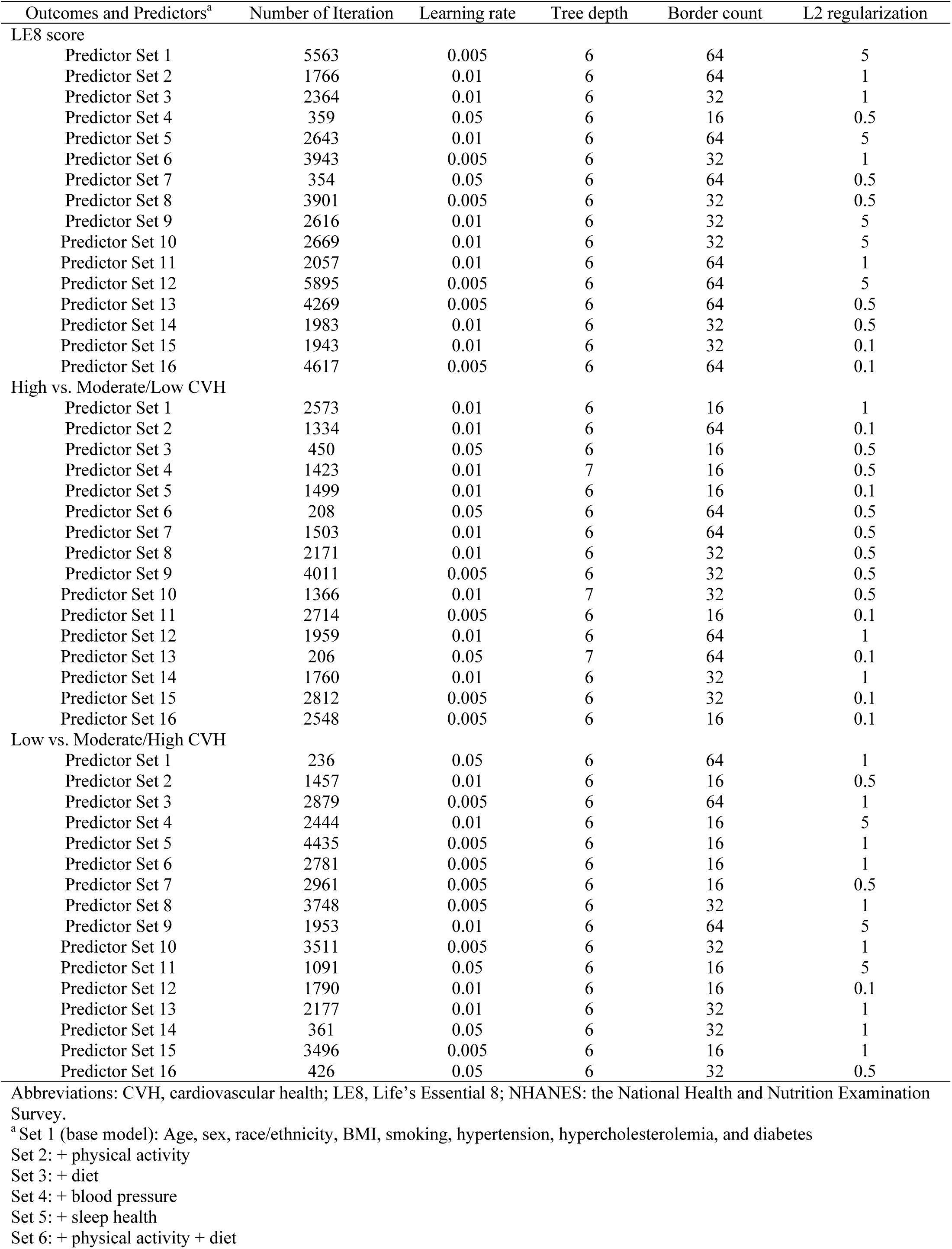
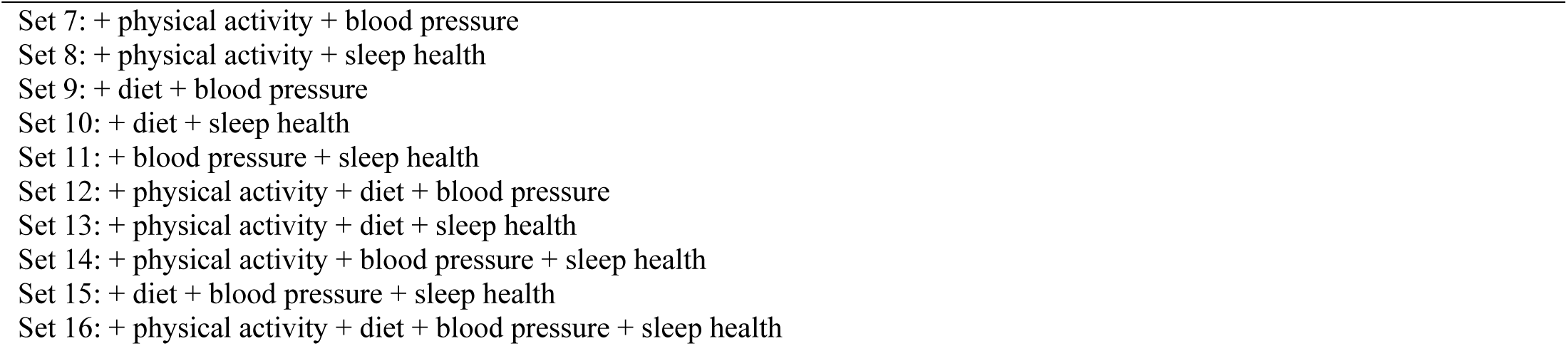
Optimal hyperparameters of predictive models of CVH based on LE8 tuned by cross-validation in the training set using the NHANES (n=27,194).

| Outcomes and Predictors<sup>a</sup> | Number of Iteration | Learning rate | Tree depth | Border count | L2 regularization |
| --- | --- | --- | --- | --- | --- |
| LE8 score |  |  |  |  |  |
| Predictor Set 1 | 5563 | 0.005 | 6 | 64 | 5 |
| Predictor Set 2 | 1766 | 0.01 | 6 | 64 | 1 |
| Predictor Set 3 | 2364 | 0.01 | 6 | 32 | 1 |
| Predictor Set 4 | 359 | 0.05 | 6 | 16 | 0.5 |
| Predictor Set 5 | 2643 | 0.01 | 6 | 64 | 5 |
| Predictor Set 6 | 3943 | 0.005 | 6 | 32 | 1 |
| Predictor Set 7 | 354 | 0.05 | 6 | 64 | 0.5 |
| Predictor Set 8 | 3901 | 0.005 | 6 | 32 | 0.5 |
| Predictor Set 9 | 2616 | 0.01 | 6 | 32 | 5 |
| Predictor Set 10 | 2669 | 0.01 | 6 | 32 | 5 |
| Predictor Set 11 | 2057 | 0.01 | 6 | 64 | 1 |
| Predictor Set 12 | 5895 | 0.005 | 6 | 64 | 5 |
| Predictor Set 13 | 4269 | 0.005 | 6 | 64 | 0.5 |
| Predictor Set 14 | 1983 | 0.01 | 6 | 32 | 0.5 |
| Predictor Set 15 | 1943 | 0.01 | 6 | 32 | 0.1 |
| Predictor Set 16 | 4617 | 0.005 | 6 | 64 | 0.1 |
| High vs. Moderate/Low CVH |  |  |  |  |  |
| Predictor Set 1 | 2573 | 0.01 | 6 | 16 | 1 |
| Predictor Set 2 | 1334 | 0.01 | 6 | 64 | 0.1 |
| Predictor Set 3 | 450 | 0.05 | 6 | 16 | 0.5 |
| Predictor Set 4 | 1423 | 0.01 | 7 | 16 | 0.5 |
| Predictor Set 5 | 1499 | 0.01 | 6 | 16 | 0.1 |
| Predictor Set 6 | 208 | 0.05 | 6 | 64 | 0.5 |
| Predictor Set 7 | 1503 | 0.01 | 6 | 64 | 0.5 |
| Predictor Set 8 | 2171 | 0.01 | 6 | 32 | 0.5 |
| Predictor Set 9 | 4011 | 0.005 | 6 | 32 | 0.5 |
| Predictor Set 10 | 1366 | 0.01 | 7 | 32 | 0.5 |
| Predictor Set 11 | 2714 | 0.005 | 6 | 16 | 0.1 |
| Predictor Set 12 | 1959 | 0.01 | 6 | 64 | 1 |
| Predictor Set 13 | 206 | 0.05 | 7 | 64 | 0.1 |
| Predictor Set 14 | 1760 | 0.01 | 6 | 32 | 1 |
| Predictor Set 15 | 2812 | 0.005 | 6 | 32 | 0.1 |
| Predictor Set 16 | 2548 | 0.005 | 6 | 16 | 0.1 |
| Low vs. Moderate/High CVH |  |  |  |  |  |
| Predictor Set 1 | 236 | 0.05 | 6 | 64 | 1 |
| Predictor Set 2 | 1457 | 0.01 | 6 | 16 | 0.5 |
| Predictor Set 3 | 2879 | 0.005 | 6 | 64 | 1 |
| Predictor Set 4 | 2444 | 0.01 | 6 | 16 | 5 |
| Predictor Set 5 | 4435 | 0.005 | 6 | 16 | 1 |
| Predictor Set 6 | 2781 | 0.005 | 6 | 16 | 1 |
| Predictor Set 7 | 2961 | 0.005 | 6 | 16 | 0.5 |
| Predictor Set 8 | 3748 | 0.005 | 6 | 32 | 1 |
| Predictor Set 9 | 1953 | 0.01 | 6 | 64 | 5 |
| Predictor Set 10 | 3511 | 0.005 | 6 | 32 | 1 |
| Predictor Set 11 | 1091 | 0.05 | 6 | 16 | 5 |
| Predictor Set 12 | 1790 | 0.01 | 6 | 16 | 0.1 |
| Predictor Set 13 | 2177 | 0.01 | 6 | 32 | 1 |
| Predictor Set 14 | 361 | 0.05 | 6 | 32 | 1 |
| Predictor Set 15 | 3496 | 0.005 | 6 | 16 | 1 |
| Predictor Set 16 | 426 | 0.05 | 6 | 32 | 0.5 |

Abbreviations: CVH, cardiovascular health; LE8, Life's Essential 8; NHANES: the National Health and Nutrition Examination Survey.
<sup>a</sup> Set 1 (base model): Age, sex, race/ethnicity, BMI, smoking, hypertension, hypercholesterolemia, and diabetes
Set 2: + physical activity
Set 3: + diet
Set 4: + blood pressure
Set 5: + sleep health
Set 6: + physical activity + diet

**Table S5.**
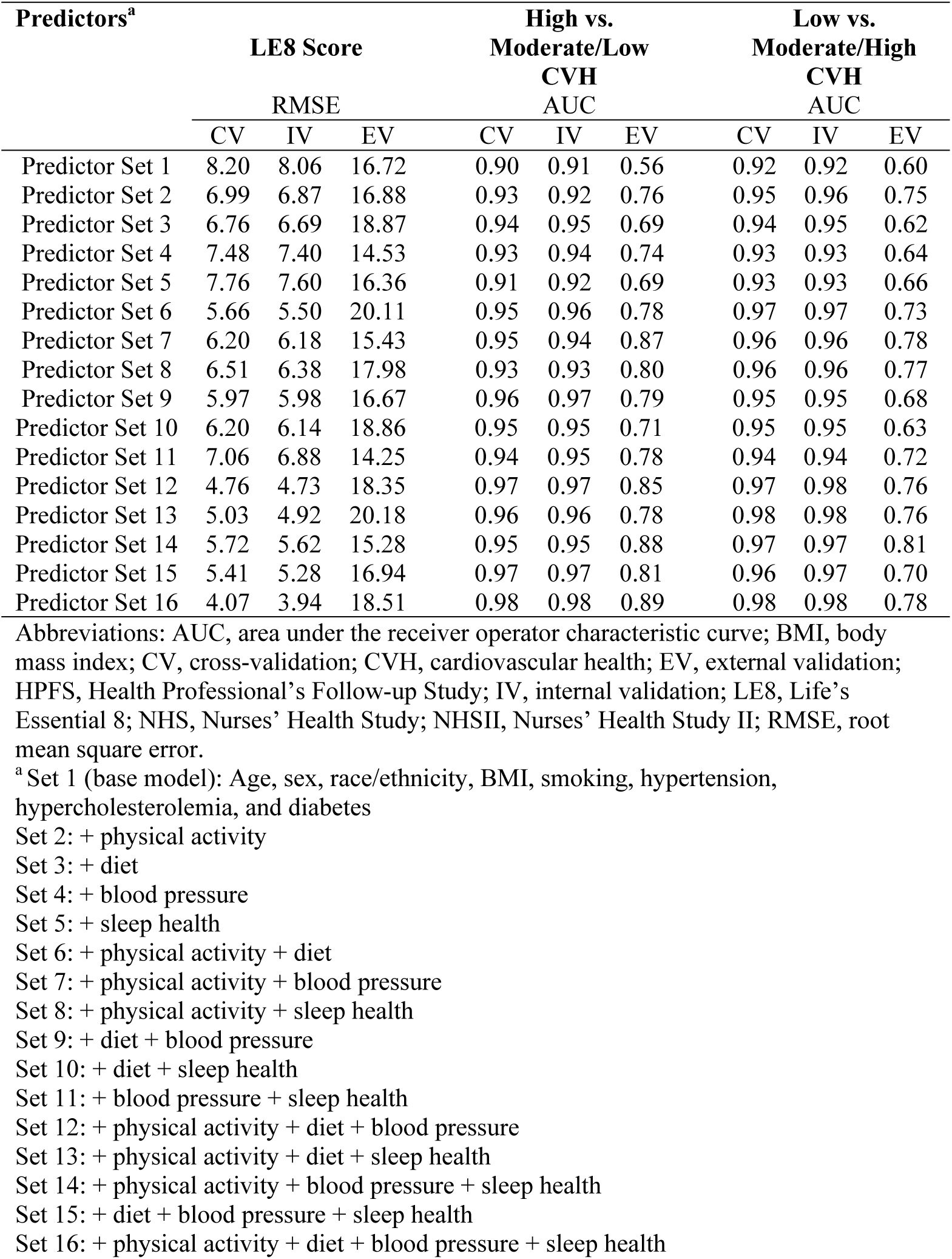
Performance of models to estimate overall CVH based on LE8 using NHS, NHSII, and HPFS (n=5,588).

**Table S6.** Performance of models to estimate overall CVH based on LE8 using the 2005-2016 NHANES (n=27,194).

| Predictors<sup>a</sup> | LE8 Score |  |  | High vs.<br>Moderate/Low<br>CVH<br>AUC |  |  | Low vs.<br>Moderate/High<br>CVH<br>AUC |  |  |
| --- | --- | --- | --- | --- | --- | --- | --- | --- | --- |
|  | RMSE |  |  |  |  |  |  |  |  |
|  | CV | IV | EV | CV | IV | EV | CV | IV | EV |
| Predictor Set 1 | 9.19 | 9.21 | 18.33 | 0.91 | 0.91 | 0.70 | 0.88 | 0.89 | 0.51 |
| Predictor Set 2 | 7.01 | 7.13 | 14.18 | 0.95 | 0.95 | 0.75 | 0.93 | 0.93 | 0.65 |
| Predictor Set 3 | 8.59 | 8.62 | 14.90 | 0.93 | 0.92 | 0.75 | 0.90 | 0.91 | 0.55 |
| Predictor Set 4 | 8.53 | 8.52 | 18.62 | 0.92 | 0.93 | 0.79 | 0.90 | 0.91 | 0.60 |
| Predictor Set 5 | 8.55 | 8.60 | 17.97 | 0.92 | 0.92 | 0.72 | 0.90 | 0.90 | 0.59 |
| Predictor Set 6 | 6.22 | 6.32 | 11.62 | 0.97 | 0.97 | 0.80 | 0.94 | 0.95 | 0.70 |
| Predictor Set 7 | 6.13 | 6.23 | 13.34 | 0.97 | 0.97 | 0.84 | 0.95 | 0.95 | 0.73 |
| Predictor Set 8 | 6.33 | 6.45 | 13.17 | 0.96 | 0.96 | 0.77 | 0.95 | 0.94 | 0.69 |
| Predictor Set 9 | 7.86 | 7.87 | 14.84 | 0.94 | 0.95 | 0.81 | 0.92 | 0.92 | 0.64 |
| Predictor Set 10 | 7.87 | 7.93 | 14.19 | 0.94 | 0.93 | 0.75 | 0.92 | 0.92 | 0.62 |
| Predictor Set 11 | 7.82 | 7.84 | 17.38 | 0.93 | 0.94 | 0.81 | 0.92 | 0.92 | 0.66 |
| Predictor Set 12 | 5.19 | 5.29 | 10.82 | 0.98 | 0.98 | 0.87 | 0.96 | 0.96 | 0.76 |
| Predictor Set 13 | 5.40 | 5.49 | 11.59 | 0.98 | 0.97 | 0.82 | 0.96 | 0.96 | 0.71 |
| Predictor Set 14 | 5.33 | 5.41 | 12.67 | 0.97 | 0.97 | 0.85 | 0.96 | 0.96 | 0.74 |
| Predictor Set 15 | 7.05 | 7.09 | 13.61 | 0.95 | 0.95 | 0.83 | 0.94 | 0.94 | 0.70 |
| Predictor Set 16 | 4.16 | 4.24 | 10.39 | 0.99 | 0.99 | 0.89 | 0.97 | 0.97 | 0.77 |

Abbreviations: AUC, area under the receiver operator characteristic curve; BMI, body mass index; CV, cross-validation; CVH, cardiovascular health; EV, external validation; LE8, Life's Essential 8; NHANES: the National Health and Nutrition Examination Survey; RMSE, root mean square error.
<sup>a</sup> Set 1 (base model): Age, sex, race/ethnicity, BMI, smoking, hypertension, hypercholesterolemia, and diabetes
Set 2: + physical activity
Set 3: + diet
Set 4: + blood pressure
Set 5: + sleep health
Set 6: + physical activity + diet
Set 7: + physical activity + blood pressure
Set 8: + physical activity + sleep health
Set 9: + diet + blood pressure
Set 10: + diet + sleep health

**Table S7.** Internal validation of models to estimate overall CVH based on LE8 in the testing sets of NHS, NHSII, and HPFS (n=5,588).

| Predictors | LE8 Score |  |  | High vs.<br>Moderate/Low CVH |  |  | Low vs.<br>Moderate/High CVH |  |  |
| --- | --- | --- | --- | --- | --- | --- | --- | --- | --- |
|  | RMSE |  |  | AUC |  |  | AUC |  |  |
|  | NHS | NHSII | HPFS | NHS | NHSII | HPFS | NHS | NHSII | HPFS |
| Predictor Set 1 | 8.21 | 8.28 | 6.94 | 0.88 | 0.92 | 0.93 | 0.92 | 0.97 | 0.91 |
| Predictor Set 2 | 6.98 | 7.01 | 6.15 | 0.90 | 0.90 | 0.86 | 0.96 | 0.97 | 0.95 |
| Predictor Set 3 | 6.85 | 6.90 | 5.55 | 0.93 | 0.93 | 0.96 | 0.94 | 0.98 | 0.93 |
| Predictor Set 4 | 7.55 | 7.50 | 6.48 | 0.92 | 0.93 | 0.98 | 0.93 | 0.98 | 0.91 |
| Predictor Set 5 | 7.77 | 7.85 | 6.41 | 0.90 | 0.93 | 0.93 | 0.93 | 0.97 | 0.94 |
| Predictor Set 6 | 5.65 | 5.68 | 4.45 | 0.95 | 0.94 | 0.94 | 0.97 | 0.98 | 0.97 |
| Predictor Set 7 | 6.27 | 6.48 | 5.38 | 0.93 | 0.93 | 0.98 | 0.96 | 0.97 | 0.95 |
| Predictor Set 8 | 6.50 | 6.44 | 5.56 | 0.91 | 0.92 | 0.93 | 0.96 | 0.98 | 0.96 |
| Predictor Set 9 | 6.13 | 5.97 | 5.17 | 0.96 | 0.97 | 0.99 | 0.95 | 0.98 | 0.94 |
| Predictor Set 10 | 6.33 | 6.24 | 4.94 | 0.94 | 0.95 | 0.94 | 0.95 | 0.97 | 0.97 |
| Predictor Set 11 | 7.00 | 7.13 | 5.93 | 0.93 | 0.94 | 0.99 | 0.94 | 0.98 | 0.95 |
| Predictor Set 12 | 4.85 | 4.85 | 3.88 | 0.97 | 0.97 | 0.99 | 0.97 | 0.98 | 0.97 |
| Predictor Set 13 | 5.12 | 4.71 | 3.91 | 0.95 | 0.96 | 0.96 | 0.97 | 0.98 | 0.98 |
| Predictor Set 14 | 5.69 | 5.94 | 4.92 | 0.94 | 0.94 | 0.99 | 0.97 | 0.98 | 0.97 |
| Predictor Set 15 | 5.44 | 5.13 | 4.47 | 0.96 | 0.98 | 0.99 | 0.96 | 0.98 | 0.97 |
| Predictor Set 16 | 4.10 | 3.65 | 3.23 | 0.98 | 0.98 | 0.99 | 0.98 | 0.98 | 0.99 |

Abbreviations: AUC, area under the receiver operator characteristic curve; BMI, body mass index; CV, cross-validation; CVH, cardiovascular health; EV, external validation; HPFS, Health Professional's Follow-up Study; IV, internal validation; LE8, Life's Essential 8; NHS, Nurses' Health Study; NHSII, Nurses' Health Study II; RMSE, root mean square error.
<sup>a</sup> Set 1 (base model): Age, gender, race/ethnicity, BMI, smoking, hypertension, hypercholesterolemia, and diabetes
Set 2: + physical activity
Set 3: + diet
Set 4: + blood pressure
Set 5: + sleep health
Set 6: + physical activity + diet
Set 7: + physical activity + blood pressure
Set 8: + physical activity + sleep health
Set 9: + diet + blood pressure
Set 10: + diet + sleep health

**Table S8.**
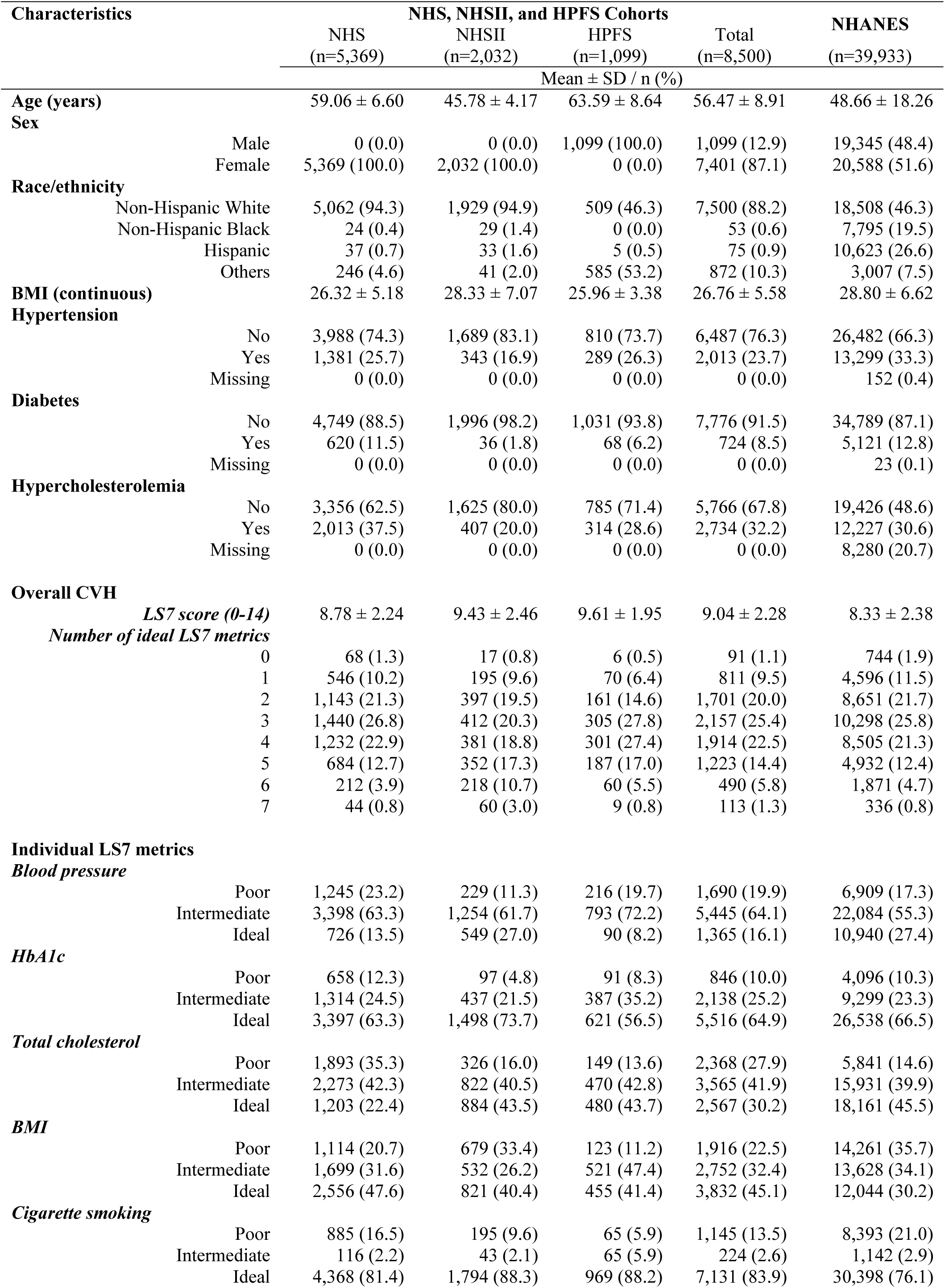
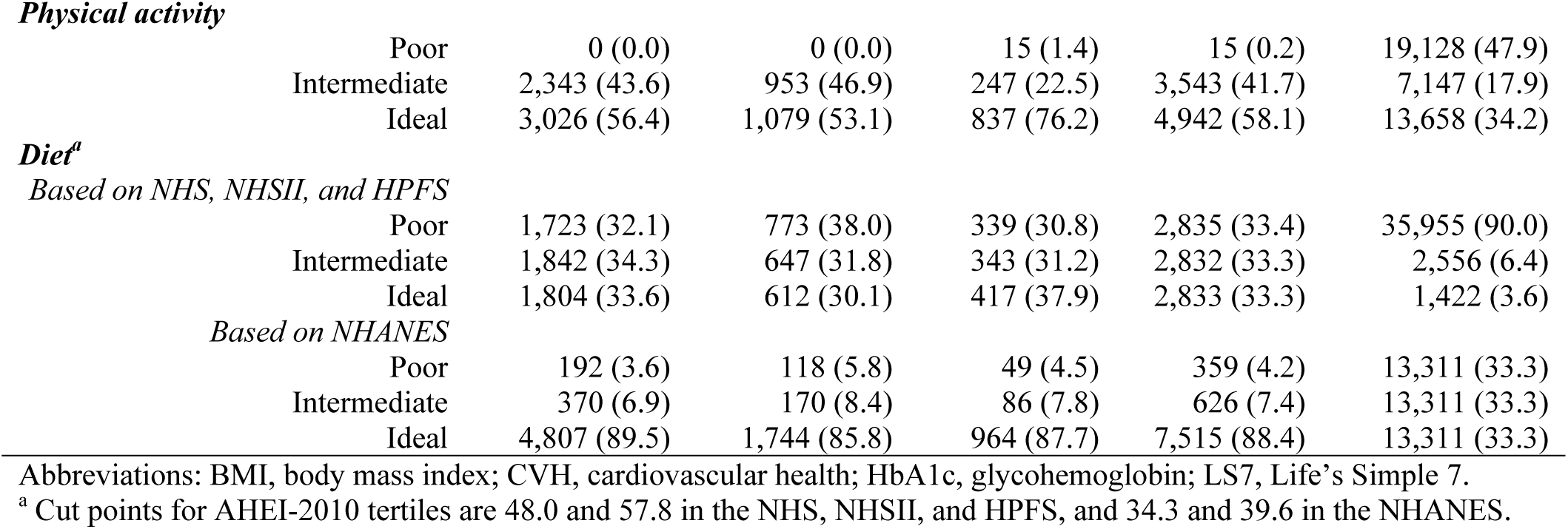
Characteristics of participants in NHS, NHSII, and HPFS, and the 1999-2016 NHANES included in developing prediction models of Life’s Simple 7 score.

| Characteristics |  | NHS, NHSII, and HPFS Cohorts |  |  |  | NHANES<br>(n=39,933) |
| --- | --- | --- | --- | --- | --- | --- |
|  |  | NHS<br>(n=5,369) | NHSII<br>(n=2,032) | HPFS<br>(n=1,099) | Total<br>(n=8,500) |  |
|  |  | Mean ± SD / n (%) |  |  |  |  |
| Age (years) |  | 59.06 ± 6.60 | 45.78 ± 4.17 | 63.59 ± 8.64 | 56.47 ± 8.91 | 48.66 ± 18.26 |
| Sex |  |  |  |  |  |  |
|  | Male | 0 (0.0) | 0 (0.0) | 1,099 (100.0) | 1,099 (12.9) | 19,345 (48.4) |
|  | Female | 5,369 (100.0) | 2,032 (100.0) | 0 (0.0) | 7,401 (87.1) | 20,588 (51.6) |
| Race/ethnicity |  |  |  |  |  |  |
|  | Non-Hispanic White | 5,062 (94.3) | 1,929 (94.9) | 509 (46.3) | 7,500 (88.2) | 18,508 (46.3) |
|  | Non-Hispanic Black | 24 (0.4) | 29 (1.4) | 0 (0.0) | 53 (0.6) | 7,795 (19.5) |
|  | Hispanic | 37 (0.7) | 33 (1.6) | 5 (0.5) | 75 (0.9) | 10,623 (26.6) |
|  | Others | 246 (4.6) | 41 (2.0) | 585 (53.2) | 872 (10.3) | 3,007 (7.5) |
| BMI (continuous) |  | 26.32 ± 5.18 | 28.33 ± 7.07 | 25.96 ± 3.38 | 26.76 ± 5.58 | 28.80 ± 6.62 |
| Hypertension |  |  |  |  |  |  |
|  | No | 3,988 (74.3) | 1,689 (83.1) | 810 (73.7) | 6,487 (76.3) | 26,482 (66.3) |
|  | Yes | 1,381 (25.7) | 343 (16.9) | 289 (26.3) | 2,013 (23.7) | 13,299 (33.3) |
|  | Missing | 0 (0.0) | 0 (0.0) | 0 (0.0) | 0 (0.0) | 152 (0.4) |
| Diabetes |  |  |  |  |  |  |
|  | No | 4,749 (88.5) | 1,996 (98.2) | 1,031 (93.8) | 7,776 (91.5) | 34,789 (87.1) |
|  | Yes | 620 (11.5) | 36 (1.8) | 68 (6.2) | 724 (8.5) | 5,121 (12.8) |
|  | Missing | 0 (0.0) | 0 (0.0) | 0 (0.0) | 0 (0.0) | 23 (0.1) |
| Hypercholesterolemia |  |  |  |  |  |  |
|  | No | 3,356 (62.5) | 1,625 (80.0) | 785 (71.4) | 5,766 (67.8) | 19,426 (48.6) |
|  | Yes | 2,013 (37.5) | 407 (20.0) | 314 (28.6) | 2,734 (32.2) | 12,227 (30.6) |
|  | Missing | 0 (0.0) | 0 (0.0) | 0 (0.0) | 0 (0.0) | 8,280 (20.7) |
| Overall CVH |  |  |  |  |  |  |
|  | LS7 score (0-14) | 8.78 ± 2.24 | 9.43 ± 2.46 | 9.61 ± 1.95 | 9.04 ± 2.28 | 8.33 ± 2.38 |
|  | Number of ideal LS7 metrics |  |  |  |  |  |
|  | 0 | 68 (1.3) | 17 (0.8) | 6 (0.5) | 91 (1.1) | 744 (1.9) |
|  | 1 | 546 (10.2) | 195 (9.6) | 70 (6.4) | 811 (9.5) | 4,596 (11.5) |
|  | 2 | 1,143 (21.3) | 397 (19.5) | 161 (14.6) | 1,701 (20.0) | 8,651 (21.7) |
|  | 3 | 1,440 (26.8) | 412 (20.3) | 305 (27.8) | 2,157 (25.4) | 10,298 (25.8) |
|  | 4 | 1,232 (22.9) | 381 (18.8) | 301 (27.4) | 1,914 (22.5) | 8,505 (21.3) |
|  | 5 | 684 (12.7) | 352 (17.3) | 187 (17.0) | 1,223 (14.4) | 4,932 (12.4) |
|  | 6 | 212 (3.9) | 218 (10.7) | 60 (5.5) | 490 (5.8) | 1,871 (4.7) |
|  | 7 | 44 (0.8) | 60 (3.0) | 9 (0.8) | 113 (1.3) | 336 (0.8) |
| Individual LS7 metrics |  |  |  |  |  |  |
| Blood pressure |  |  |  |  |  |  |
|  | Poor | 1,245 (23.2) | 229 (11.3) | 216 (19.7) | 1,690 (19.9) | 6,909 (17.3) |
|  | Intermediate | 3,398 (63.3) | 1,254 (61.7) | 793 (72.2) | 5,445 (64.1) | 22,084 (55.3) |
|  | Ideal | 726 (13.5) | 549 (27.0) | 90 (8.2) | 1,365 (16.1) | 10,940 (27.4) |
| HbA1c |  |  |  |  |  |  |
|  | Poor | 658 (12.3) | 97 (4.8) | 91 (8.3) | 846 (10.0) | 4,096 (10.3) |
|  | Intermediate | 1,314 (24.5) | 437 (21.5) | 387 (35.2) | 2,138 (25.2) | 9,299 (23.3) |
|  | Ideal | 3,397 (63.3) | 1,498 (73.7) | 621 (56.5) | 5,516 (64.9) | 26,538 (66.5) |
| Total cholesterol |  |  |  |  |  |  |
|  | Poor | 1,893 (35.3) | 326 (16.0) | 149 (13.6) | 2,368 (27.9) | 5,841 (14.6) |
|  | Intermediate | 2,273 (42.3) | 822 (40.5) | 470 (42.8) | 3,565 (41.9) | 15,931 (39.9) |
|  | Ideal | 1,203 (22.4) | 884 (43.5) | 480 (43.7) | 2,567 (30.2) | 18,161 (45.5) |
| BMI |  |  |  |  |  |  |
|  | Poor | 1,114 (20.7) | 679 (33.4) | 123 (11.2) | 1,916 (22.5) | 14,261 (35.7) |
|  | Intermediate | 1,699 (31.6) | 532 (26.2) | 521 (47.4) | 2,752 (32.4) | 13,628 (34.1) |
|  | Ideal | 2,556 (47.6) | 821 (40.4) | 455 (41.4) | 3,832 (45.1) | 12,044 (30.2) |
| Cigarette smoking |  |  |  |  |  |  |
|  | Poor | 885 (16.5) | 195 (9.6) | 65 (5.9) | 1,145 (13.5) | 8,393 (21.0) |
|  | Intermediate | 116 (2.2) | 43 (2.1) | 65 (5.9) | 224 (2.6) | 1,142 (2.9) |
|  | Ideal | 4,368 (81.4) | 1,794 (88.3) | 969 (88.2) | 7,131 (83.9) | 30,398 (76.1) |

**Physical activity**
|  |  |  |  |  |  |
| --- | --- | --- | --- | --- | --- |
| Poor | 0 (0.0) | 0 (0.0) | 15 (1.4) | 15 (0.2) | 19,128 (47.9) |
| Intermediate | 2,343 (43.6) | 953 (46.9) | 247 (22.5) | 3,543 (41.7) | 7,147 (17.9) |
| Ideal | 3,026 (56.4) | 1,079 (53.1) | 837 (76.2) | 4,942 (58.1) | 13,658 (34.2) |

**Diet<sup>a</sup>***Based on NHS, NHSII, and HPFS*
|  |  |  |  |  |  |
| --- | --- | --- | --- | --- | --- |
| Poor | 1,723 (32.1) | 773 (38.0) | 339 (30.8) | 2,835 (33.4) | 35,955 (90.0) |
| Intermediate | 1,842 (34.3) | 647 (31.8) | 343 (31.2) | 2,832 (33.3) | 2,556 (6.4) |
| Ideal | 1,804 (33.6) | 612 (30.1) | 417 (37.9) | 2,833 (33.3) | 1,422 (3.6) |

*Based on NHANES*
|  |  |  |  |  |  |
| --- | --- | --- | --- | --- | --- |
| Poor | 192 (3.6) | 118 (5.8) | 49 (4.5) | 359 (4.2) | 13,311 (33.3) |
| Intermediate | 370 (6.9) | 170 (8.4) | 86 (7.8) | 626 (7.4) | 13,311 (33.3) |
| Ideal | 4,807 (89.5) | 1,744 (85.8) | 964 (87.7) | 7,515 (88.4) | 13,311 (33.3) |

Abbreviations: BMI, body mass index; CVH, cardiovascular health; HbA1c, glycohemoglobin; LS7, Life's Simple 7.
<sup>a</sup> Cut points for AHEI-2010 tertiles are 48.0 and 57.8 in the NHS, NHSII, and HPFS, and 34.3 and 39.6 in the NHANES.

**Table S9.** Optimal hyperparameters of predictive models of CVH based on LS7 tuned by cross-validation in the training set using NHS, NHSII, and HPFS (n=8,500).

| Outcomes and Predictors<sup>a</sup> | Number of Iteration | Learning rate | Tree depth | Border count | L2 regularization |
| --- | --- | --- | --- | --- | --- |
| $\geq 1$ Ideal CVH metrics | | | | | |
| Predictor Set 1 | 569 | 0.05 | 6 | 32 | 5 |
| Predictor Set 2 | 683 | 0.05 | 6 | 64 | 0.1 |
| Predictor Set 3 | 743 | 0.05 | 6 | 32 | 1 |
| Predictor Set 4 | 623 | 0.01 | 6 | 16 | 0.5 |
| Predictor Set 5 | 483 | 0.05 | 6 | 32 | 1 |
| Predictor Set 6 | 269 | 0.01 | 7 | 16 | 0.1 |
| Predictor Set 7 | 82 | 0.05 | 7 | 32 | 0.1 |
| Predictor Set 8 | 303 | 0.05 | 6 | 16 | 0.5 |
| $\geq 2$ Ideal CVH metrics | | | | | |
| Predictor Set 1 | 88 | 0.05 | 7 | 64 | 1 |
| Predictor Set 2 | 220 | 0.05 | 6 | 64 | 0.5 |
| Predictor Set 3 | 477 | 0.05 | 6 | 64 | 5 |
| Predictor Set 4 | 107 | 0.05 | 7 | 32 | 0.5 |
| Predictor Set 5 | 285 | 0.05 | 6 | 64 | 0.1 |
| Predictor Set 6 | 315 | 0.05 | 6 | 16 | 1 |
| Predictor Set 7 | 422 | 0.05 | 6 | 64 | 1 |
| Predictor Set 8 | 326 | 0.05 | 6 | 16 | 0.5 |
| $\geq 3$ Ideal CVH metrics | | | | | |
| Predictor Set 1 | 1017 | 0.01 | 6 | 64 | 0.5 |
| Predictor Set 2 | 279 | 0.05 | 6 | 64 | 5 |
| Predictor Set 3 | 1273 | 0.01 | 6 | 16 | 0.1 |
| Predictor Set 4 | 1552 | 0.01 | 6 | 32 | 1 |
| Predictor Set 5 | 2942 | 0.005 | 6 | 32 | 1 |
| Predictor Set 6 | 1901 | 0.005 | 6 | 32 | 5 |
| Predictor Set 7 | 1310 | 0.01 | 6 | 64 | 0.1 |
| Predictor Set 8 | 1091 | 0.01 | 6 | 16 | 0.5 |
| $\geq 4$ Ideal CVH metrics | | | | | |
| Predictor Set 1 | 128 | 0.05 | 6 | 32 | 1 |
| Predictor Set 2 | 136 | 0.05 | 6 | 32 | 1 |
| Predictor Set 3 | 297 | 0.05 | 6 | 64 | 1 |
| Predictor Set 4 | 113 | 0.05 | 6 | 64 | 1 |
| Predictor Set 5 | 293 | 0.05 | 6 | 16 | 0.5 |
| Predictor Set 6 | 108 | 0.05 | 6 | 32 | 1 |
| Predictor Set 7 | 2225 | 0.01 | 6 | 64 | 5 |
| Predictor Set 8 | 1965 | 0.01 | 6 | 16 | 1 |
| $\geq 5$ Ideal CVH metrics | | | | | |
| Predictor Set 1 | 259 | 0.05 | 6 | 16 | 0.5 |
| Predictor Set 2 | 1064 | 0.01 | 6 | 16 | 0.1 |
| Predictor Set 3 | 1394 | 0.01 | 7 | 32 | 0.1 |
| Predictor Set 4 | 168 | 0.05 | 6 | 64 | 1 |
| Predictor Set 5 | 2088 | 0.01 | 6 | 16 | 0.1 |
| Predictor Set 6 | 952 | 0.01 | 6 | 16 | 0.5 |
| Predictor Set 7 | 2461 | 0.005 | 6 | 32 | 0.1 |
| Predictor Set 8 | 2421 | 0.005 | 6 | 64 | 1 |
| $\geq 6$ Ideal CVH metrics | | | | | |
| Predictor Set 1 | 229 | 0.05 | 6 | 32 | 0.5 |
| Predictor Set 2 | 134 | 0.05 | 7 | 32 | 0.1 |
| Predictor Set 3 | 2086 | 0.05 | 6 | 16 | 5 |
| Predictor Set 4 | 378 | 0.05 | 6 | 16 | 0.5 |
| Predictor Set 5 | 184 | 0.05 | 7 | 64 | 0.1 |
| Predictor Set 6 | 162 | 0.05 | 7 | 64 | 0.5 |
| Predictor Set 7 | 108 | 0.05 | 7 | 16 | 0.5 |
| Predictor Set 8 | 1511 | 0.01 | 6 | 64 | 0.5 |
| 7 Ideal CVH metrics |  |  |  |  |  |
| Predictor Set 1 | 183 | 0.05 | 7 | 32 | 1 |
| Predictor Set 2 | 192 | 0.05 | 7 | 32 | 0.1 |
| Predictor Set 3 | 179 | 0.05 | 6 | 32 | 0.5 |
| Predictor Set 4 | 87 | 0.05 | 6 | 64 | 1 |
| Predictor Set 5 | 167 | 0.05 | 7 | 32 | 0.1 |
| Predictor Set 6 | 110 | 0.05 | 6 | 64 | 1 |
| Predictor Set 7 | 252 | 0.05 | 7 | 64 | 0.5 |
| Predictor Set 8 | 66 | 0.05 | 8 | 32 | 0.1 |
| LS7 score |  |  |  |  |  |
| Predictor Set 1 | 1702 | 0.01 | 6 | 64 | 5 |
| Predictor Set 2 | 241 | 0.05 | 8 | 16 | 5 |
| Predictor Set 3 | 4406 | 0.005 | 6 | 32 | 5 |
| Predictor Set 4 | 213 | 0.05 | 7 | 16 | 5 |
| Predictor Set 5 | 2033 | 0.01 | 6 | 64 | 5 |
| Predictor Set 6 | 1177 | 0.01 | 6 | 32 | 1 |
| Predictor Set 7 | 3436 | 0.005 | 6 | 64 | 5 |
| Predictor Set 8 | 2409 | 0.01 | 6 | 64 | 1 |

Abbreviations: AHEI-2010: alternative healthy eating index 2010; CVH, cardiovascular health; HPFS, Health Professional's Follow-up Study; LS7, Life's Simple 7; NHS, Nurses' Health Study; NHSII, Nurses' Health Study II.
<sup>a</sup>Set 1 (base model): Age, gender, race/ethnicity, BMI, smoking, hypertension, hypercholesterolemia, and diabetes
Set 2: base model + physical activity
Set 3: base model + diet (AHEI-2010)
Set 4: base model + blood pressure
Set 5: base model + physical activity + diet (AHEI-2010)
Set 6: base model + physical activity + blood pressure
Set 7: base model + diet (AHEI-2010) + blood pressure
Set 8: base model + physical activity + diet (AHEI-2010) + blood pressure

**Table S10.** Optimal hyperparameters of predictive models of CVH based on LS7 tuned by cross-validation in the training set using the NHANES (n=39,933).

| Outcomes and Predictors<sup>a</sup> | Number of Iteration | Learning rate | Tree depth | Border count | L2 regularization |
| --- | --- | --- | --- | --- | --- |
| $\geq 1$ Ideal CVH metrics | | | | | |
| Predictor Set 1 | 93 | 0.05 | 7 | 32 | 0.1 |
| Predictor Set 2 | 100 | 0.05 | 6 | 64 | 0.1 |
| Predictor Set 3 | 713 | 0.01 | 7 | 64 | 1 |
| Predictor Set 4 | 116 | 0.05 | 8 | 64 | 0.1 |
| Predictor Set 5 | 917 | 0.005 | 7 | 64 | 0.5 |
| Predictor Set 6 | 911 | 0.005 | 8 | 64 | 0.1 |
| Predictor Set 7 | 1311 | 0.01 | 8 | 64 | 1 |
| Predictor Set 8 | 452 | 0.01 | 8 | 64 | 0.5 |
| $\geq 2$ Ideal CVH metrics | | | | | |
| Predictor Set 1 | 3693 | 0.005 | 6 | 32 | 0.1 |
| Predictor Set 2 | 3819 | 0.005 | 6 | 32 | 0.1 |
| Predictor Set 3 | 342 | 0.05 | 6 | 32 | 0.5 |
| Predictor Set 4 | 2113 | 0.01 | 6 | 32 | 0.1 |
| Predictor Set 5 | 391 | 0.05 | 6 | 64 | 0.5 |
| Predictor Set 6 | 1911 | 0.01 | 6 | 32 | 0.5 |
| Predictor Set 7 | 1669 | 0.01 | 7 | 32 | 0.1 |
| Predictor Set 8 | 4081 | 0.005 | 7 | 32 | 1 |
| $\geq 3$ Ideal CVH metrics | | | | | |
| Predictor Set 1 | 1692 | 0.01 | 6 | 64 | 0.5 |
| Predictor Set 2 | 2323 | 0.01 | 6 | 32 | 1 |
| Predictor Set 3 | 1063 | 0.05 | 6 | 32 | 5 |
| Predictor Set 4 | 3895 | 0.005 | 6 | 64 | 1 |
| Predictor Set 5 | 2630 | 0.01 | 6 | 64 | 0.5 |
| Predictor Set 6 | 3526 | 0.005 | 7 | 32 | 1 |
| Predictor Set 7 | 2403 | 0.01 | 6 | 64 | 0.5 |
| Predictor Set 8 | 311 | 0.05 | 7 | 64 | 0.5 |
| $\geq 4$ Ideal CVH metrics | | | | | |
| Predictor Set 1 | 5189 | 0.005 | 6 | 32 | 1 |
| Predictor Set 2 | 4550 | 0.005 | 6 | 64 | 1 |
| Predictor Set 3 | 2305 | 0.01 | 6 | 64 | 1 |
| Predictor Set 4 | 3138 | 0.01 | 6 | 32 | 1 |
| Predictor Set 5 | 3628 | 0.005 | 6 | 64 | 0.1 |
| Predictor Set 6 | 383 | 0.05 | 6 | 32 | 1 |
| Predictor Set 7 | 4408 | 0.005 | 6 | 64 | 1 |
| Predictor Set 8 | 2585 | 0.01 | 7 | 32 | 1 |
| $\geq 5$ Ideal CVH metrics | | | | | |
| Predictor Set 1 | 651 | 0.05 | 7 | 64 | 5 |
| Predictor Set 2 | 339 | 0.05 | 8 | 32 | 5 |
| Predictor Set 3 | 3245 | 0.01 | 8 | 64 | 5 |
| Predictor Set 4 | 204 | 0.05 | 7 | 64 | 0.5 |
| Predictor Set 5 | 3850 | 0.005 | 6 | 64 | 0.1 |
| Predictor Set 6 | 415 | 0.05 | 8 | 64 | 5 |
| Predictor Set 7 | 371 | 0.05 | 6 | 64 | 0.5 |
| Predictor Set 8 | 887 | 0.05 | 6 | 64 | 5 |
| $\geq 6$ Ideal CVH metrics | | | | | |
| Predictor Set 1 | 1638 | 0.01 | 6 | 32 | 0.1 |
| Predictor Set 2 | 1179 | 0.01 | 6 | 32 | 0.1 |
| Predictor Set 3 | 1247 | 0.01 | 6 | 32 | 0.5 |
| Predictor Set 4 | 339 | 0.05 | 6 | 16 | 0.1 |
| Predictor Set 5 | 245 | 0.05 | 7 | 32 | 0.5 |
| Predictor Set 6 | 1909 | 0.01 | 7 | 16 | 0.1 |
| Predictor Set 7 | 258 | 0.05 | 6 | 16 | 0.1 |
| Predictor Set 8 | 1559 | 0.01 | 6 | 16 | 0.5 |
| 7 Ideal CVH metrics |  |  |  |  |  |
| Predictor Set 1 | 382 | 0.05 | 7 | 16 | 0.5 |
| Predictor Set 2 | 562 | 0.05 | 7 | 16 | 1 |
| Predictor Set 3 | 243 | 0.05 | 7 | 16 | 0.5 |
| Predictor Set 4 | 391 | 0.05 | 7 | 16 | 0.5 |
| Predictor Set 5 | 188 | 0.05 | 8 | 16 | 0.1 |
| Predictor Set 6 | 298 | 0.05 | 6 | 16 | 0.1 |
| Predictor Set 7 | 208 | 0.05 | 8 | 32 | 1 |
| Predictor Set 8 | 210 | 0.05 | 8 | 16 | 0.5 |
| LS7 score |  |  |  |  |  |
| Predictor Set 1 | 551 | 0.05 | 6 | 32 | 0.5 |
| Predictor Set 2 | 850 | 0.05 | 6 | 64 | 5 |
| Predictor Set 3 | 6403 | 0.005 | 6 | 64 | 5 |
| Predictor Set 4 | 544 | 0.05 | 6 | 32 | 1 |
| Predictor Set 5 | 831 | 0.05 | 6 | 32 | 5 |
| Predictor Set 6 | 3534 | 0.01 | 6 | 32 | 5 |
| Predictor Set 7 | 6494 | 0.005 | 6 | 16 | 1 |
| Predictor Set 8 | 3301 | 0.01 | 6 | 32 | 0.5 |

Abbreviations: CVH, cardiovascular health; LS7, Life's Simple 7; NHANES: the National Health and Nutrition Examination Survey.
<sup>a</sup>Set 1 (base model): Age, sex, race/ethnicity, BMI, smoking, hypertension, hypercholesterolemia, and diabetes
Set 2: + physical activity
Set 3: + diet
Set 4: + blood pressure
Set 5: + physical activity + diet
Set 6: + physical activity + blood pressure
Set 7: + diet + blood pressure
Set 8: + physical activity + diet + blood pressure

**Table S11.**
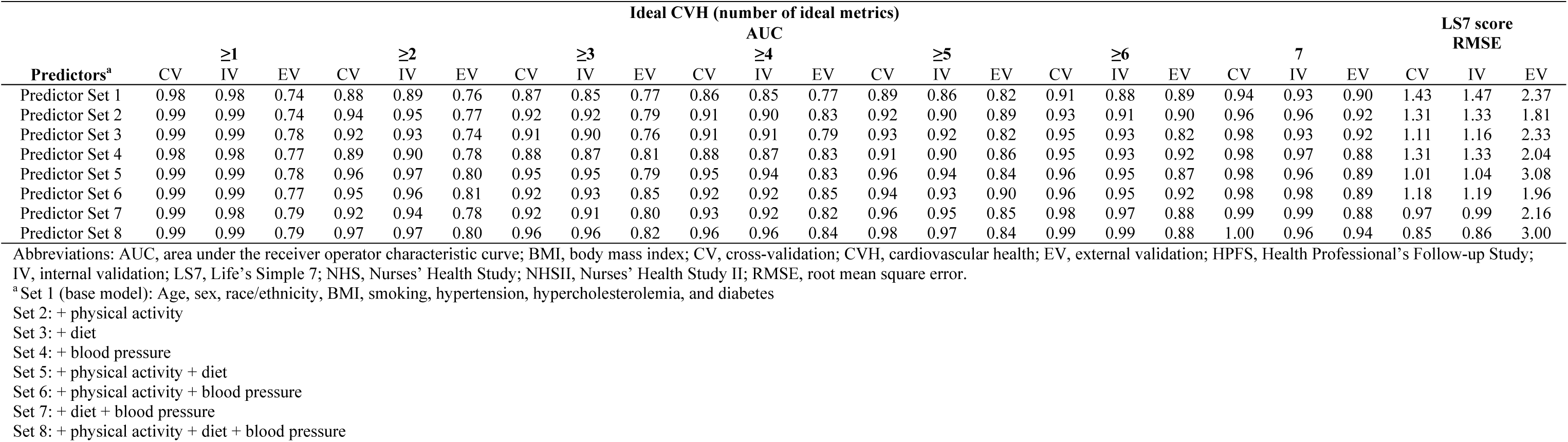
Performance of models to estimate overall CVH based on LS7 using NHS, NHSII, and HPFS (n=8,500).

**Table S12.**
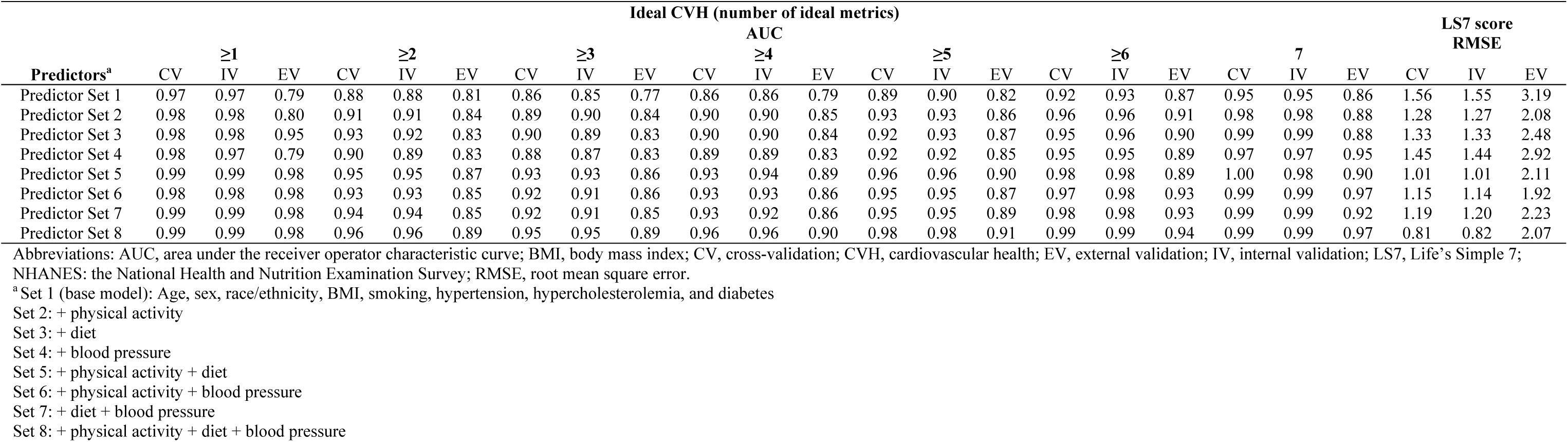
Performance of models to estimate overall CVH based on LS7 using the 1999-2016 NHANES (n=39,933).

**Table S13.**
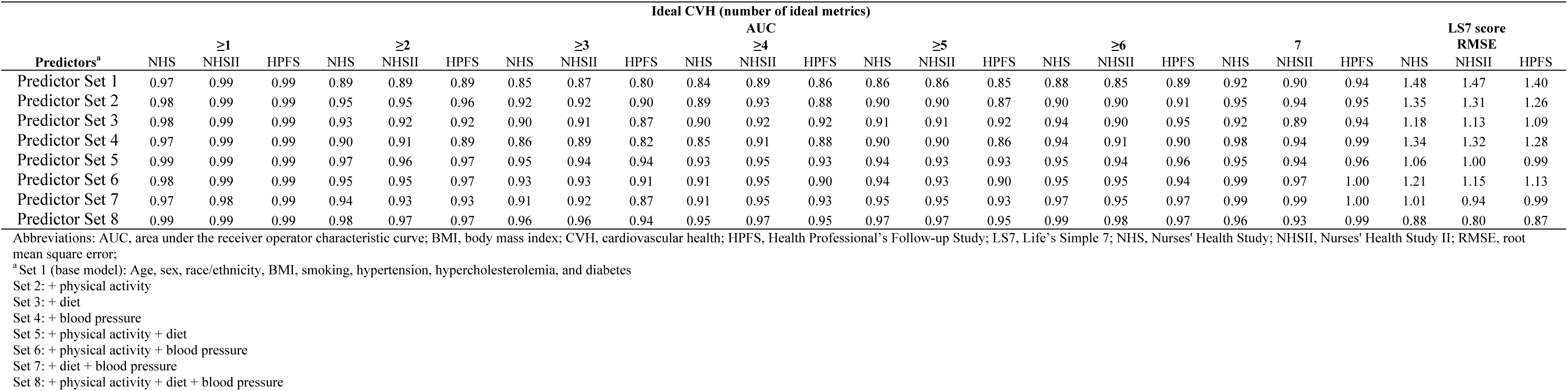
Internal validation of models to estimate overall CVH based on LS7 in the testing sets of NHS, NHSII, and HPFS (n=8,500).

**Figure S1.**
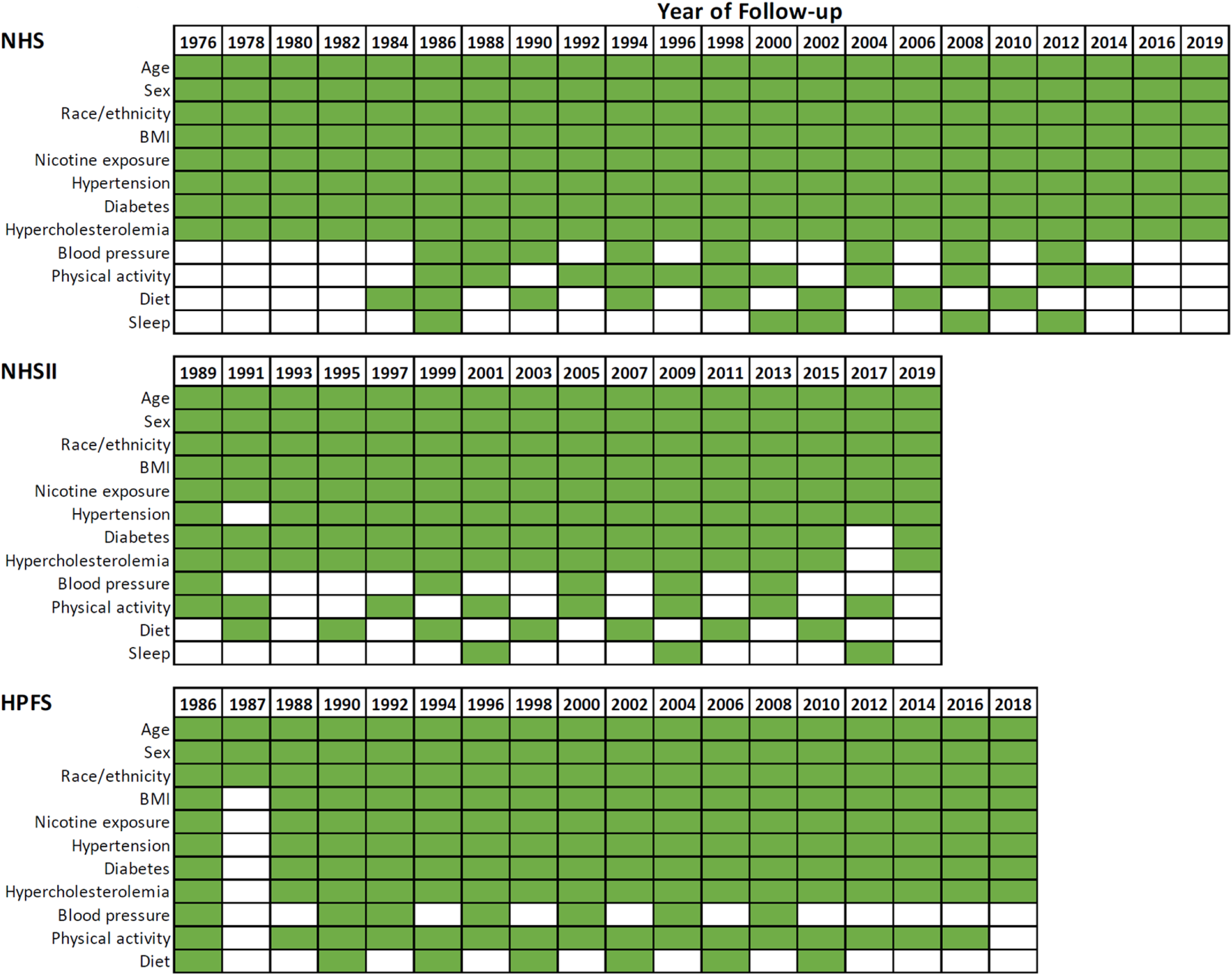
Availabilities of CVH metrics and predictors in NHS, NHSII, and HPFS. Abbreviations: BMI, body mass index; HPFS, Health Professional’s Follow-up Study; NHS, Nurses’ Health Study; NHSII, Nurses’ Health Study II.

**Figure S2.**
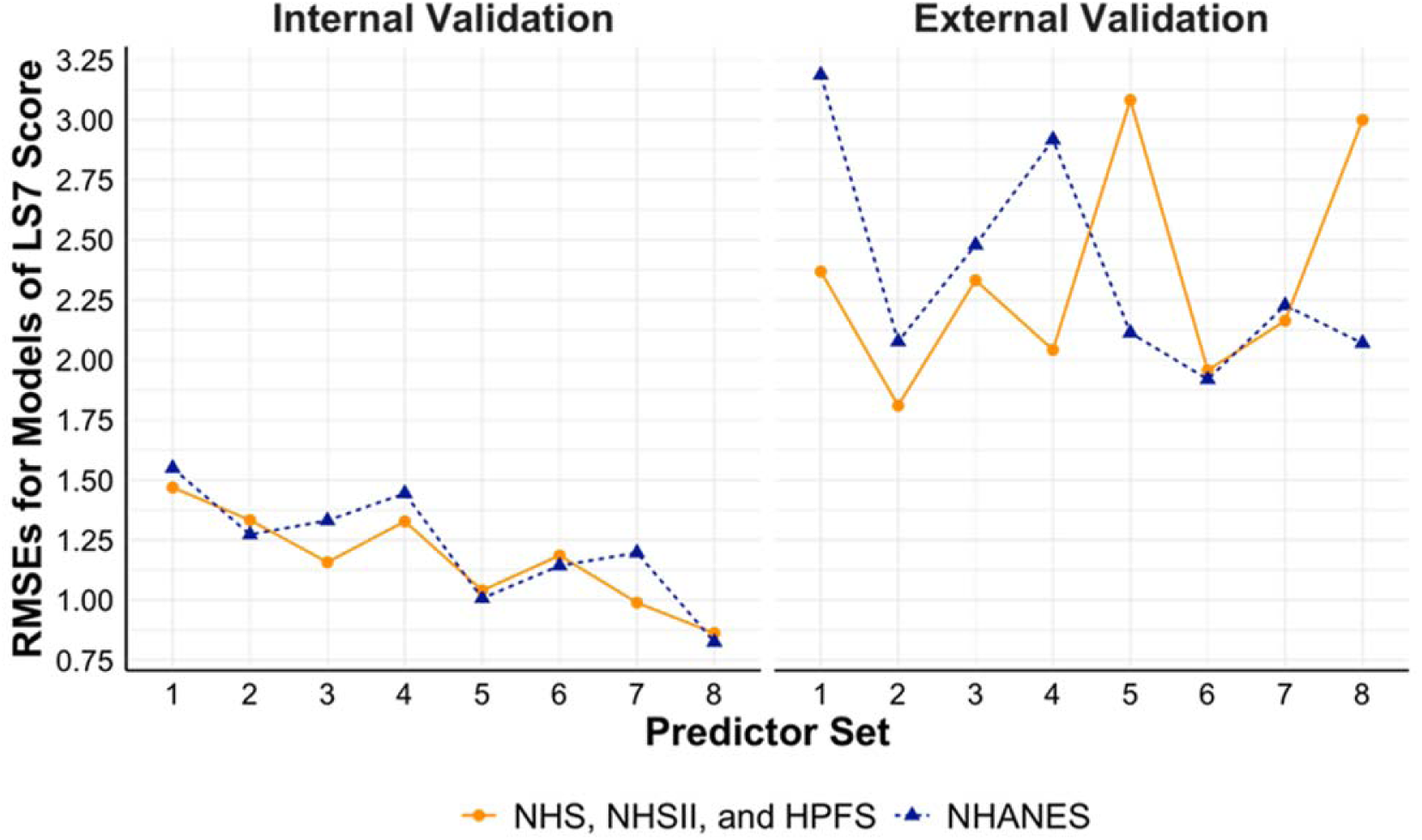
Performance of models to estimate continuous overall CVH score based on LS7 using NHS, NHSII, and HPFS (n=8,500), and NHANES (n=39,933). Set 1 (base model): Age, sex, race/ethnicity, BMI, smoking, hypertension, hypercholesterolemia, and diabetes Set 2: + physical activity Set 3: + diet Set 4: + blood pressure Set 5: + physical activity + diet Set 6: + physical activity + blood pressure Set 7: + diet + blood pressure Set 8: + physical activity + diet + blood pressure Abbreviations: BMI, body mass index; CVH, cardiovascular health; HPFS, Health Professional’s Follow-up Study; LS7, Life’s Simple 7; NHANES: the National Health and Nutrition Examination Survey; NHS, Nurses’ Health Study; NHSII, Nurses’ Health Study II; RMSE, root mean square error.

**Figure S3.**
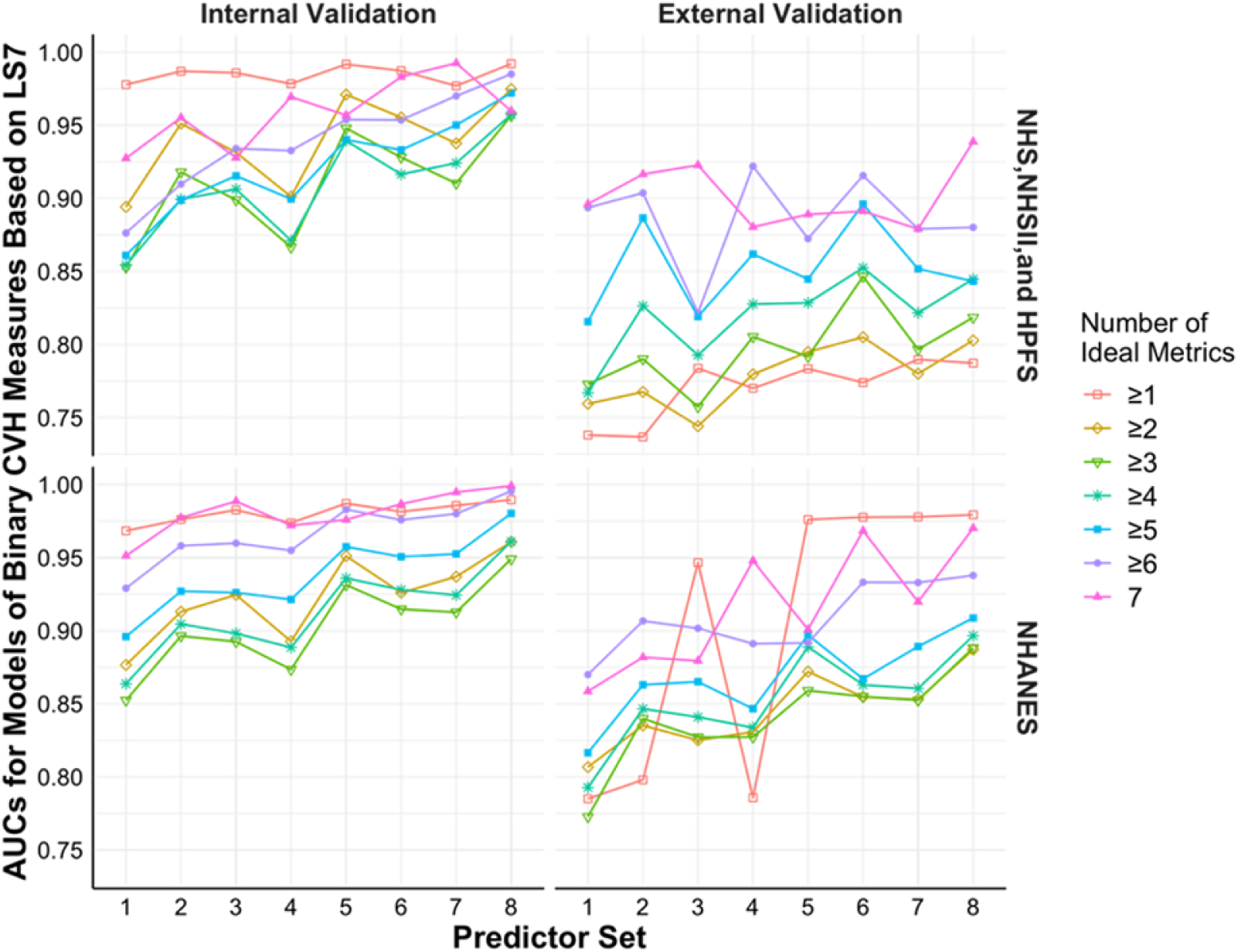
Performance of models to estimate binary CVH measures based on LS7 using NHS, NHSII, and HPFS (n=8,500), and NHANES (n=39,933). Set 1 (base model): Age, sex, race/ethnicity, BMI, smoking, hypertension, hypercholesterolemia, and diabetes; Set 2: + physical activity Set 3: + diet Set 4: + blood pressure Set 5: + physical activity + diet Set 6: + physical activity + blood pressure Set 7: + diet + blood pressure Set 8: + physical activity + diet + blood pressure Abbreviations: AUC, area under the receiver operator characteristic curve; BMI, body mass index; CVH, cardiovascular health; HPFS, Health Professional’s Follow-up Study; LS7, Life’s Simple 7; NHANES: the National Health and Nutrition Examination Survey; NHS, Nurses’ Health Study; NHSII, Nurses’ Health Study II.

**Figure S4.**
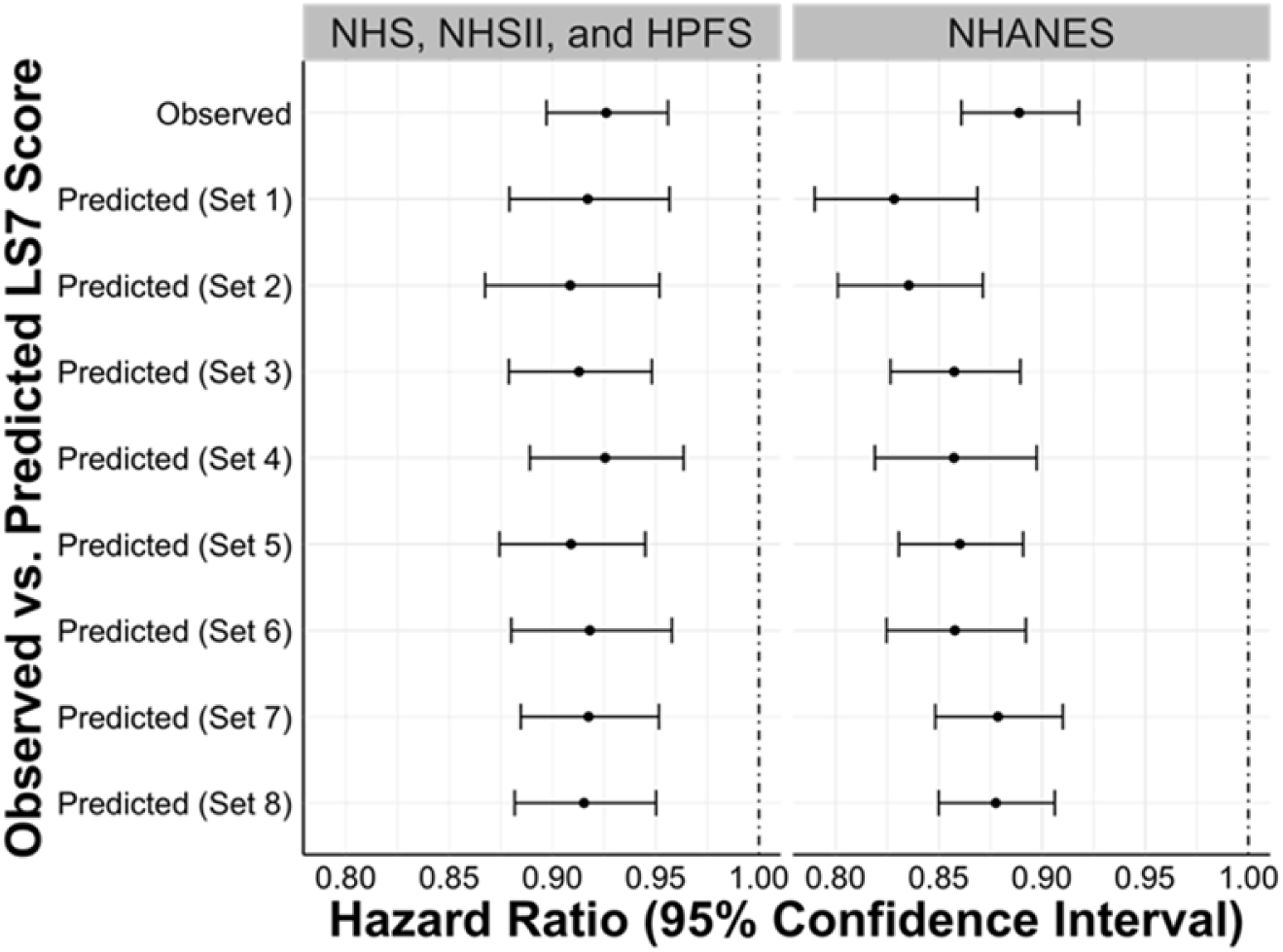
Hazard ratios (95% confidence intervals) of all-cause mortality with original vs. predicted overall CVH score based on LS7 in testing sets of NHS, NHSII, and HPFS (n=8,500), and NHANES (n=39,933). Set 1 (base model): Age, sex, race/ethnicity, BMI, smoking, hypertension, hypercholesterolemia, and diabetes; Set 2: + physical activity Set 3: + diet Set 4: + blood pressure Set 5: + physical activity + diet Set 6: + physical activity + blood pressure Set 7: + diet + blood pressure Set 8: + physical activity + diet + blood pressure Abbreviations: BMI, body mass index; CVH, cardiovascular health; HPFS, Health Professional’s Follow-up Study; LS7, Life’s Simple 7; NHANES: the National Health and Nutrition Examination Survey; NHS, Nurses’ Health Study; NHSII, Nurses’ Health Study II.

